## Supplemental File for "Maternal prenatal anxiety and depression and trajectories of cardiometabolic risk factors across childhood and adolescence: a prospective cohort study"

**List of contents**

**Supplemental Methods**

Supplemental Methods S1 Details of study exposure measurement and prevalence

Supplemental Methods S2 Details of measurement sources for cardiometabolic risk factors

Supplemental Methods S3 Details on measurement of confounders

Supplemental Methods S4 Details of model selection for outcomes

**Supplemental Tables**

Supplemental Table S1 Model details for log BMI trajectories

Supplemental Table S2 Model details for log fat mass trajectories

Supplemental Table S3 Model details for lean mass trajectories

Supplemental Table S4 Model details for SBP, DBP and pulse rate trajectories

Supplemental Table S5 Model details for glucose trajectories

Supplemental Table S6 Model details for log insulin and triglyceride trajectories

Supplemental Table S7 Model details for HDL-c and non-HDL-c trajectories

Supplemental Table S8 Number of participants with cardiometabolic measures at each time point

Supplemental Table S9 Characteristics at birth of the mothers of children included in models compared with those excluded due to missing exposure, outcome or confounder data

Supplemental Table S10 Mean trajectories of BMI estimated from multilevel models, by maternal anxiety during pregnancy

Supplemental Table S11 Mean trajectories of fat mass estimated from multilevel models, by maternal anxiety during pregnancy

Supplemental Table S12 Mean trajectories of blood pressure and pulse rate estimated from multilevel models, by maternal anxiety during pregnancy

Supplemental Table S13 Mean trajectories of lean mass estimated from multilevel models, by maternal anxiety during pregnancy

Supplemental Table S14 Mean trajectories of glucose estimated from multilevel models, by maternal anxiety during pregnancy

Supplemental Table S15 Mean trajectories of log insulin and triglyceride estimated from multilevel models, by maternal anxiety during pregnancy

Supplemental Table S16 Mean trajectories of HDL-c and non-HDL-c estimated from multilevel models, by maternal anxiety during pregnancy

Supplemental Table S17 Mean trajectories of BMI estimated from multilevel models, by maternal depression during pregnancy

Supplemental Table S18 Mean trajectories of fat mass estimated from multilevel models, by maternal depression during pregnancy

Supplemental Table S19 Mean trajectories of blood pressure and pulse rate estimated from multilevel models, by maternal depression during pregnancy

Supplemental Table S20 Mean trajectories of lean mass estimated from multilevel models, by maternal depression during pregnancy

Supplemental Table S21 Mean trajectories of glucose estimated from multilevel models, by maternal depression during pregnancy

Supplemental Table S22 Mean trajectories of log insulin and triglyceride estimated from multilevel models, by maternal depression during pregnancy

Supplemental Table S23 Mean trajectories of HDL-c and non-HDL-estimated from multilevel models, by maternal depression during pregnancy

**Supplemental Figures**

Supplemental Figure S1 Flow diagram of study design

Supplemental Figure S2 Mean predicted confounder adjusted trajectories of lean mass (9 to 18 years), by continuous maternal anxiety levels during pregnancy

Supplemental Figure S3 Mean predicted confounder adjusted trajectories of log fat mass (9 to 18 years), by continuous maternal anxiety levels during pregnancy

Supplemental Figure S4 Mean predicted confounder adjusted trajectories of log BMI (1 to 18 years), by continuous maternal anxiety levels during pregnancy

Supplemental Figure S5 Mean predicted confounder adjusted trajectories of pulse rate from 7 to 18 years by continuous maternal anxiety levels during pregnancy

Supplemental Figure S6 Mean predicted confounder adjusted trajectories of SBP from 7 to 18 years by continuous maternal anxiety levels during pregnancy

Supplemental Figure S7 Mean predicted confounder adjusted trajectories of DBP from 7 to 18 years by continuous maternal anxiety levels during pregnancy

Supplemental Figure S8 Mean predicted confounder adjusted trajectories of log insulin from (birth to 18 years) by continuous maternal anxiety levels during pregnancy

Supplemental Figure S9 Mean predicted confounder adjusted trajectories of glucose (7 to18 years), by continuous maternal anxiety levels during pregnancy

Supplemental Figure S10 Mean predicted confounder adjusted trajectories of HDL-c (birth to18 years), by continuous maternal anxiety levels during pregnancy

Supplemental Figure S11 Mean predicted confounder adjusted trajectories of triglyceride (birth to18 years), by continuous maternal anxiety levels during pregnancy

Supplemental Figure S12 Mean predicted confounder adjusted trajectories of non-HDL-c (birth to18 years), by continuous maternal anxiety levels during pregnancy

Supplemental Figure S13 Mean predicted confounder adjusted trajectories of lean mass (9 to 18 years), log fat mass (9 to 18 years) and log BMI (1 to 18 years), by categorical maternal depression levels during pregnancy

Supplemental Figure S14 Mean predicted confounder adjusted trajectories of pulse rate, SBP and DBP from 7 to 18 years, by categorical maternal depression levels during pregnancy

Supplemental Figure S15 Mean predicted confounder adjusted trajectories of log insulin (birth to 18 years) and glucose (7 to18 years), by categorical maternal depression levels during pregnancy

Supplemental Figure S16 Mean predicted confounder adjusted trajectories of HDL-c, triglyceride and non-HDL-c from birth to 18 years, by categorical maternal depression levels during pregnancy

Supplemental Figure S17 Mean predicted confounder adjusted trajectories of lean mass (9 to 18 years), by continuous maternal depression levels during pregnancy

Supplemental Figure S18 Mean predicted confounder adjusted trajectories of log fat mass (9 to 18 years), by continuous maternal depression levels during pregnancy

Supplemental Figure S19 Mean predicted confounder adjusted trajectories of log BMI (1 to 18 years), by continuous maternal depression levels during pregnancy

Supplemental Figure S20 Mean predicted confounder adjusted trajectories of pulse rate from 7 to 18 years by continuous maternal depression levels during pregnancy

Supplemental Figure S21 Mean predicted confounder adjusted trajectories of SBP from 7 to 18 years by continuous maternal depression levels during pregnancy

Supplemental Figure S22 Mean predicted confounder adjusted trajectories of DBP from 7 to 18 years by continuous maternal depression levels during pregnancy

Supplemental Figure S23 Mean predicted confounder adjusted trajectories of log insulin from (birth to 18 years) by continuous maternal depression levels during pregnancy

Supplemental Figure S24 Mean predicted confounder adjusted trajectories of glucose (7 to18 years), by continuous maternal depression levels during pregnancy

Supplemental Figure S25 Mean predicted confounder adjusted trajectories of HDL-c (birth to18 years), by continuous maternal depression levels during pregnancy

Supplemental Figure S26 Mean predicted confounder adjusted trajectories of triglyceride (birth to18 years), by continuous maternal depression levels during pregnancy

Supplemental Figure S27 Mean predicted confounder adjusted trajectories of non-HDL-c (birth to18 years), by continuous maternal depression levels during pregnancy

**Supplemental Methods S1 Details of study exposure measurement and prevalence**

***Maternal self-report prenatal anxiety***

Maternal prenatal anxiety was measured using the eight items from the anxiety subscale of Crown Crisp Experiential Index (CCEI). The CCEI is a validated self-report measure, with anxiety items rated on a 4-point scale (“very often” to “never”); a higher score is indicative of higher anxiety. Items ask women how they feel ‘at this stage in pregnancy’ and include “Do you sometimes feel panicky”. This scale has previously demonstrated internal consistency coefficients above 0.80 in the ALSPAC cohort during pregnancy^8^.

***Maternal self-report depression***

Prenatal depression was measured using the Edinburgh Postnatal Depression Scale (EPDS). This is a validated 10-item self-report measure, designed and used to screen women for depression before and after pregnancy. The EPDS has an internal consistency coefficient of .87^2^. Items ask women to rate how they felt in the last week in relation to statements such as, ‘Things have been getting on top of me’. A higher score is indicative of greater depressive symptoms, with a score of 13 and above considered indicative of depression; the cut-off of 13 is typically used to determine presence of clinical depression^33^.

**Supplemental Methods S2 Details of measurement sources for cardiometabolic risk factors**

***Details on measurement of height and weight at research clinics***

Data from age 1 onwards are included in this analysis. We did not include measures before 1 year because of the difficulty in accurately modelling BMI from birth through the whole of childhood due to its early peak followed by adiposity rebound. From 1 to 5 years, measures were available from routine child health clinics for most children and extracted from health visitor records, which form part of standard child care in the UK. Data were also available from research clinic measurements on a random 10% subsample of the cohort. All cohort members were invited to research clinics from age 7 onwards. Across all ages parent-reported measures were available.

At the clinics, crown-heel length for children aged four to 25 months was measured using a Harpenden Neonatometer and from 25 months onwards standing height was measured using a Leicester Height Measure; weight was measured using Fereday 100kg combined scale (four-month clinic), Soenhle scale or Seca scale model 724 (eight-month clinic), Seca 724 or Seca 835 (12-month clinic), Seca 835 (18 months onwards). From age 7 years, all children were invited to annual clinics, at which standing height was measured to the last complete mm using the Harpenden Stadiometer and weight was measured to the nearest 0.1kg using the Tanita Body Fat Analyser (Model TBF 305).

***Details on measurement of blood pressure***

A Dinamap 9301 Vital Signs Monitor (Morton Medical, London) was used at 7, 9, 11, 15 and 18 years; an Omron MI-5 was used at the 10-year clinic; a Dinamap 8100 Vital Signs Monitor (Morton Medical) was used at the 12-year clinic.

***Details on measurement of blood based biomarkers***

Non-fasting glucose was measured at age 7 years using Nuclear Magnetic Resonance (NMR) spectroscopy. In a random 10% of the cohort at age 9 years, fasting glucose and insulin were also available; these were taken as part of a continuation of an earlier sub-study called “Child in Focus” that included approximately 10% of the overall cohort. Fasting glucose and insulin were available from research clinics held when participants were 15 and 18 years old^54, 55^.

Plasma lipid assays (triglyceride and high-density lipoprotein cholesterol (HDL-c)) were performed by modification of the standard Lipid Research Clinics Protocol using enzymatic reagents for lipid determination. All assay coefficients of variation were <5%. Samples were collected after an overnight fast and were analysed by the hexokinase method. Insulin was measured by an ELISA (Mercodia, Uppsala, Sweden) that does not cross-react with proinsulin. All assay coefficients of variation were <5%.

***Details on measurement on biomarkers using Nuclear Magnetic Resonance (NMR) spectroscopy***

A comprehensive profiling of offspring circulating lipids, lipoproteins, and metabolites was done by a high-throughput NMR metabolomics platform, providing a snapshot of offspring serum metabolome. At age 7, this was done on fasted blood samples and glucose is included in our analyses. At age 15 and 18, non-fasted bloods were used.

**Supplemental Methods S3 Details on measurement of confounders**

Data on confounders was collected using self-report measures at 18 and/or 32 weeks gestation.

Household social class was measured as the highest of the mother’s or her partner’s occupational social class using data on job title and details of occupation collected about the mother and her partner from the mother’s questionnaire at 32 weeks gestation. Social class was derived using the standard occupational classification (SOC) codes developed by the United Kingdom Office of Population Census and Surveys and classified as I professional, II managerial and technical, IIINM non-manual, IIIM manual, and IV&V part skilled occupations and unskilled occupation.

Parity data was collected using a self-report item asking ‘How many times have you been pregnant altogether before this time?’, with responses categorised as 0, 1 or 2 based on participant responses.

Maternal age at delivery was collected as self-report data and measured in years.

Maternal Education maternal education was reported at 32 weeks gestation according to increasing levels of achievement. These levels were: less than an Ordinary level (O Level), which was categorised as no education or a certificate of secondary education, subject-specific qualifications at a lower level than O levels that were obtained by age 16 years; O level, which are subject-specific qualifications generally obtained at age 16 years; Advanced level (A Level), which are subject-specific qualifications generally obtained at age 18 years; and university degree or above.

Maternal smoking during pregnancy was self-reported in response to the question “How many times per day did you smoke” at 18 weeks; and “How many cigarettes per day are you yourself smoking at the moment” at 32 weeks. Prenatal smoking was categorised as No smoking or Yes smoking in the current study based on responses to these questions.

Maternal pre-pregnancy BMI data was collected using a self-report measure at 12 weeks gestation. Women were asked “What was your weight before you started this pregnancy? (please indicate whether stones, pounds or kilos)” and “How tall are you? (Please indicate whether feet, inches or metres)”. BMI was calculated as kg/m^2^.

**Supplemental Methods S4 Details of model selection for outcomes**

Two approaches, fractional polynomials and linear splines were used in the modelling of trajectories as described previously.

Fractional polynomials were used for BMI, due to the complex pattern of change in BMI during childhood and adolescence. Fractional polynomials involve raising age to many combinations of powers, resulting in a wide range of possible curves and offering more flexibility than standard polynomial approaches.

Linear splines were used to model all other outcomes, as too few measurement occasions were available to permit modelling using fractional polynomials or other age term combinations. Linear splines allow knot points to be fitted at different ages to derive periods of change that are approximately linear. Models were derived by initially examining observed data for each risk factor. We also plotted mean values for each risk factor on each measurement occasion to assist on decisions regarding knot points. We compared observed and predicted measurements for a selection of suitable models for each risk factor. We examined rates of change between time periods in order to examine whether changes between periods were similar or different. In cases where rates of change between two spline periods appeared identical, the fit of models with reduced splines was explored. We also compared model fit statistics (Akaike’s Information Criterion) for several models with different knot points (with knot points placed at whole years closest to mean age at clinics due to a greater density of measures). Systolic blood pressure (SBP) and diastolic blood pressure (DBP) models have been modelled previously and are described elsewhere in detail.

In all models, age (in years) was centred at the first available measure. For each risk factor except insulin, we included all participants with at least one measure of the risk factor in each multilevel model, under a missing at random (MAR) assumption, to minimise selection bias. The observations of participants who reported being pregnant at the 18-year clinic were excluded from the multilevel models at that time point only (n=6). Models for insulin included participants with at least one measure before and after 11 years of age to improve model fit due to the sparsity of measures at the earlier time points (birth and 9 years).

**BMI** has been modelled previously using fractional polynomials and is described elsewhere^34^. BMI was modelled from age 1 as BMI is not considered an appropriate measure of adiposity in infants. Briefly, BMI was log transformed due to skewness of the data and fractional polynomials were used where age was raised to various combinations of powers (each of the following single powers, plus each combination of two powers: 0.5, 1, 2, 3, -0.5, -1, -2, natural log), from which we selected the best fitting curve (the one with the lowest likelihood value). The resulting curve contained three age terms including log age, log age* age and log age *age^^2.^ To account for the likely reduced accuracy of parent-reported measurements, a binary indicator of measurement source (research clinic or health records versus parent-report) was included as a fixed effect. The variance of measurement occasion-level residuals (the differences between observed and predicted measurements) was allowed to vary with age for log BMI. The model took the form of: log BMI_ij_ = (β_0_+u_0j_+e_0ij_) + (β_1_+u_1j_)(ln(age)_ij_) + (β_2_+u_2j_)(age*ln(age)_ij_) + (β_3_+u_3j_)(age^2^*ln(age)_ij_) + (β_8_+e_1ij_)(measurement_source_ij_) + e_ij_(age_months_ij_) where for person j at measurement occasion i; β’s represent fixed effect coefficients, u_0j_ to u_3j_ indicate person-specific random effects for the intercept and linear, quadratic and cubic age terms respectively, and e_1_ represents the occasion-specific residuals or measurement error which was allowed to vary with age and according to measurement source.

**Fat mass** and **lean mass** were measured on 5 occasions between 9 and 18 years. Fat mass was log transformed due to skewness of the data. Knots were placed at 13 and 15 resulting in three periods of change; from 9-13, 13-15, 15-18. Both models were adjusted for a time-and sex-varying height co-variate which was included as a fixed effect For lean mass, the covariance between the second spline from 13 to 15 and third spline from 15 to 18 was set to zero to improve model convergence. The models took the form of: log fat mass_ij_ / lean mass_ij_ = β_0_ + u_0j_ + (β_1_+ u_1j_ )s_ij1_ + (β_2_+ u_2j_ )s_ij2_ + (β_3_ + u_3j )_s_ij3_ + β_4_ (age&sex adjusted height covariate)_ij_ + e_ij_ where for person j at measurement occasion i; β_0_ represents the fixed effect coefficient for the average intercept, β_1_ to β_3_ represent fixed effect coefficients for the average linear slopes of each linear spline, β_4_ represents the fixed effect coefficient for the average difference in measurements between individuals of different heights, u_0j_ to u_3j_ indicate person-specific random effects for the intercept and slopes respectively, and e_ij_ represents the occasion-specific residuals or measurement error which was allowed to vary with age.

**SBP, DBP** and **pulse rate** were measured at 7 time points from 7 to 18 years^35-38^. The knots for all models were placed at 12 and 16 resulting in three periods of change; from 7 to 12, 12-16 and 16-18. All models included a fixed effect to adjust for the use of the use of an Omron MI-5 machine to measure SBP in 10-year clinic which differed from all other clinics and a binary time indicator as a level 1 random effect of age less than or greater than 10 years to account for changing measurement error with age. The models took the form of: SBP_ij_ /DBP_ij_ /pulse_ij_ = β_0_ + u_0j_ + (β_1_+ u_1j_ )s_ij1_ + (β_2_+ u_2j_ )s_ij2_ + (β_3_ + u_3j )_s_ij3_ + β_4_ (machine)_ij_ + e_ij_(age_binary_ij_) where for person j at measurement occasion i; β_0_ represents the fixed effect coefficient for the average intercept, β_1_ to β_3_ represent fixed effect coefficients for the average linear slopes of each linear spline, β_4_ represents the fixed effect coefficient for the average difference in measurements between the machine used at the 10 year clinic compared to the machine used at other clinics, u_0j_ to u_3j_ indicate person-specific random effects for the intercept and slopes respectively, and e_ij_ represents the occasion-specific residuals or measurement error which was allowed to vary with age.

**Triglyceride** and **HDL-c** were measured 5 times from birth to 18 years. Triglyceride was log transformed due to the skewness of the data. **Non-HDL-c** was derived by subtracting HDL-c from total cholesterol. Knots for triglyceride and non-HDL-c were placed at 9 and 15 years resulting in two periods of change; from birth to 9 years and 9 to 18. Knots were placed at age 7 and 15 years for HDL-c resulting in two periods of change; from birth to 7 years and 7 - 18 years. The models took the form of: log triglyceride_ij_/HDL-c_ij_/non-HDL-c_ij_ = β_0_ + u_0j_ + (β_1_+ u_1j_ )s_ij1_ + (β_2_+ u_2j_ )s_ij2_ + e_ij_ where for person j at measurement occasion i; β_0_ represents the fixed effect coefficient for the average intercept, β_1_ and β_2_ represent fixed effect coefficients for the average linear slopes of each linear spline, u_0j_ to u_3j_ indicate person-specific random effects for the intercept and slopes respectively, and e_ij_ represents the occasion-specific residuals or measurement error.

**Glucose** was measured on four occasions (7, 9, 15, and 18). A knot was placed at 15 resulting in two periods of change; from 7 to 15 and 15 to 18. Due to few available repeated measures of glucose, we modelled the person-specific random effects as a single linear slope rather than a function of the splines as was done in all other linear spline models. This allowed person specific variation from the average trajectory but under the assumption that person-specific deviation from the mean trajectory was constant over time. The model took the form of: glucose_ij_ = β_0_ + u_0j_ + (β_1_)s_ij1_ + (β_2_)s_ij2_ + u_1j_*age + e_ij_ where for person j at measurement occasion i; β_0_ represents the fixed effect coefficient for the average intercept, β_1_ and β_2_ represent fixed effect coefficients for the average linear slopes of each linear spline, u_0j_ to u_1j_ indicate person-specific random effects for the intercept and slope respectively, and e_ij_ represents the occasion-specific residuals or measurement error.

**Insulin** was measured on four occasions (birth, 9, 15, and 18). Due to the sparsity of measures at birth and 9 years, the model for insulin was restricted to participants with at least one measure before and after age 11, to improve model fit. Knots were placed at 9 and 15 resulting in three periods of change; from birth to 9 years, 9 to 15 and 15 to 18. Insulin was log transformed due to skewness of the data. The model took the form of: log insulin_ij_ = β_0_ + u_0j_ + (β_1_+ u_1j_ )s_ij1_ + (β_2_+ u_2j_ )s_ij2_ + (β_3_ + u_3j)_)s_ij3+_ e_ij_ where for person j at measurement occasion i; β_0_ represents the fixed effect coefficient for the average intercept, β_1_ to β_3_ represent fixed effect coefficients for the average linear slopes of each linear spline, u_0j_ to u_3j_ indicate person-specific random effects for the intercept and slopes respectively, and e_ij_ represents the occasion-specific residuals or measurement error.

|  | No of contributing individuals | | Assessment of model fit | | | |
| --- | --- | --- | --- | --- | --- | --- |
|  | Total number of observations | Number of individuals with 1 measure | Mean observed  BMI, log (kg/m^2^) (SD) ^a^ | Mean predicted  BMI, log (kg/m^2^) (SD) ^a^ | Mean difference (observed – predicted), log (kg/m^2^) ^a^ | 95% level of agreement between observed and predicted, log (kg/m^2^) ^a^ |
| Overall | 80796 | 8606 |  |  |  |  |
| 1-3 years | 11042 | 7131 | 2.84 (0.09) | 2.84 (0.06) | -0.01 | -0.17 to 0.15 |
| 3-7 years | 21216 | 7734 | 2.78 (0.10) | 2.79 (0.07) | -0.01 | -0.17 to 0.15 |
| 7-9 years | 7033 | 6077 | 2.77 (0.12) | 2.80 (0.10) | -0.03 | -0.17 to 0.10 |
| 9-11 years | 9850 | 5828 | 2.84 (0.14) | 2.85 (0.12) | -0.01 | -0.12 to 0.10 |
| 11-13 years | 11069 | 5318 | 2.90 (0.16) | 2.90 (0.13) | 0.0002 | -0.11 to 0.12 |
| 13-15 years | 12118 | 5312 | 2.97 (0.16) | 2.97 (0.14) | 0.0002 | -0.13 to 0.13 |
| 15-18 years | 8468 | 4518 | 3.07 (0.16) | 3.09 (0.15) | -0.02 | -0.14 to 0.10 |

**Supplemental Table S1 Model details for log BMI trajectories**

BMI, Body Mass Index; SD, standard deviation

^a^ BMI is natural log transformed. All values are in log form.

**Supplemental Table S2 Model details for log fat mass trajectories**

|  | No of contributing individuals | | Assessment of model fit | | | |
| --- | --- | --- | --- | --- | --- | --- |
|  | Total number of observations | Number of individuals with 1 measure | Mean observed fat mass, log (kg) (SD) ^a^ | Mean predicted fat mass, log (kg) (SD)^a^ | Mean difference (observed – predicted), log (kg)^a^ | 95% level of agreement between observed and predicted, log (kg)^a^ |
| Overall | 21615 | 6032 |  |  |  |  |
| 9 years | 5134 | 5134 | 1.97 (0.57) | 1.97 (0.54) | -0.01 | -0.18 to 0.17 |
| 9-13 years | 10099 | 10099 | 2.13 (0.59) | 2.12 (0.56) | 0.01 | -0.20 to 0.21 |
| 13-15 years | 4371 | 4371 | 2.44 (0.59) | 2.46 (0.55) | -0.02 | -0.28 to 0.24 |
| 15-18 years | 7145 | 7145 | 2.63 (0.60) | 2.63 (0.57) | 0.01 | -0.19 to 0.20 |

SD, standard deviation

^a^ Fat mass is natural log transformed. All values are in log form.

**Supplemental Table S3 Model details for lean mass trajectories**

|  | | No of contributing individuals | | Assessment of model fit | | | | | | |
| --- | --- | --- | --- | --- | --- | --- | --- | --- | --- | --- |
|  | | Total number of observations | Number of individuals with 1 measure | Mean observed lean mass, kg (SD) | | Mean predicted lean mass, kg (SD) | | Mean difference (observed – predicted), kg | | 95% level of agreement between observed and predicted, kg |
| Overall | 21615 | | 6032 |  |  | |  | |  | |
| 9 years | 5134 | | 5134 | 24.56 (3.16) | 24.57 (2.81) | | -0.01 | | -2.53 to 2.52 | |
| 9-13 years | 10099 | | 10099 | 27.10 (4.57) | 27.11 (4.27) | | 0.00 | | -2.45 to 2.44 | |
| 13-15 years | 4371 | | 4371 | 38.01 (6.41) | 38.01 (5.97) | | 0.00 | | -2.94 to 2.95 | |
| 15-18 years | 7145 | | 7145 | 44.35 (9.26) | 44.35 (9.06) | | 0.003 | | -1.92 to 1.92 | |

SD, standard deviation

**Supplemental Table S4 Model details for SBP, DBP and pulse rate trajectories**

|  | No of contributing individuals | | Assessment of model fit | | | |
| --- | --- | --- | --- | --- | --- | --- |
|  | Total number of observations | Number of individuals with 1 measure | Mean observed SBP, DBP or pulse rate, (SD) ^a^ | Mean predicted SBP, DBP or pulse rate, (SD) ^a^ | Mean difference (observed – predicted) ^a^ | 95% level of agreement between observed and predicted ^a^ |
| SBP |  |  |  |  |  |  |
| Overall | 32900 | 6671 |  |  |  |  |
| 7 years | 5672 | 5672 | 98.76 (9.13) | 98.62 (5.37) | 0.15 | -10.70 to 11.00 |
| 7-12 years | 20670 | 6479 | 102.50 (9.50) | 102.70 (6.21) | -0.20 | -11.80 to 11.40 |
| 12-16 years | 8694 | 5090 | 115.82 (11.77) | 115.35 (8.84) | 0.46 | -11.31 to 12.24 |
| 16-18 years | 3536 | 3428 | 116.82 (10.15) | 117.14 (7.44) | -0.31 | -12.48 to 11.85 |
| DBP |  |  |  |  |  |  |
| Overall | 32900 | 6671 |  |  |  |  |
| 7 years | 5672 | 5672 | 56.34 (6.55) | 56.56 (3.37) | -0.22 | -8.90 to 8.46 |
| 7-12 years | 20670 | 6479 | 57.99 (6.92) | 56.88 (3.39) | 1.11 | -9.09 to 11.31 |
| 12-16 years | 8694 | 5090 | 61.07 (9.61) | 61.92 (5.07) | -0.86 | -12.62 to 10.91 |
| 16-18 years | 3536 | 3428 | 64.17 (6.06) | 64.49 (3.68) | -0.32 | -13.14 to 12.50 |
| Pulse |  |  |  |  |  |  |
| Overall | 32900 | 6671 |  |  |  |  |
| 7 years | 5672 | 5672 | 83.18 (10.65) | 83.15 (6.11) | 0.03 | -13.08 to 13.13 |
| 7-12 years | 20670 | 6479 | 77.57 (11.54) | 78.91 (7.01) | -1.34 | -15.93 to 13.25 |
| 12-16 years | 8694 | 5090 | 74.00 (11.29) | 74.08 (7.03) | -0.08 | -13.88 to 13.72 |
| 16-18 years | 3536 | 3428 | 65.59 (10.04) | 65.85 (6.29) | -0.27 | -14.69 to 14.16 |

DBP, diastolic blood pressure; SBP, systolic blood pressure; SD, standard deviation.

^a^Units are in mmHg for SBP and DBP and bpm for pulse rate.

**Supplemental Table S5 Model details for glucose trajectories**

|  | No of contributing individuals | | Assessment of model fit | | | |
| --- | --- | --- | --- | --- | --- | --- |
|  | Total number of observations | Number of individuals with 1 measure | Mean observed glucose, mmol/l (SD) | Mean predicted glucose, mmol/l (SD) | Mean difference (observed – predicted), mmol/l | 95% level of agreement between observed and predicted, mmol/l |
| Overall | 9390 | 5045 |  |  |  |  |
| 7 years | 3907 | 3907 | 4.18 (0.50) | 4.22 (0.24) | -0.03 | -0.59 to 0.53 |
| 7-15 years | 4576 | 4065 | 4.29 (0.55) | 4.28 (0.27) | 0.01 | -0.59 to 0.61 |
| 15-18 years | 4812 | 3203 | 5.12 (0.39) | 5.13 (0.17) | -0.01 | -0.65 to 0.62 |

Mmol/l, millimoles per litre; SD, standard deviation

**Supplemental Table S6 Model details for log insulin and log triglyceride trajectories**

|  | No of contributing individuals | | Assessment of model fit | | | |
| --- | --- | --- | --- | --- | --- | --- |
|  | Total number of observations | Number of individuals with 1 measure | Mean observed triglycerides,  (SD) ^a^ | Mean predicted triglycerides,  (SD) ^a^ | Mean difference (observed – predicted)^a^ | 95% level of agreement between observed and predicted |
| Log insulin |  |  |  |  |  |  |
| Overall | 1464 | 526 |  |  |  |  |
| Birth | 206 | 206 | 1.05 (0.48) | 1.04 (0.17) | 0.01 | -0.63 to 0.64 |
| 0-9 years | 206 | 206 | 1.05 (0.48) | 1.04 (0.17) | 0.01 | -0.63 to 0.64 |
| 9-15 years | 445 | 443 | 1.58 (0.62) | 1.56 (0.39) | 0.02 | -0.48 to 0.52 |
| 9-18 years | 813 | 523 | 2.07 (0.53) | 2.07 (0.27) | 0.0001 | -0.63 to 0.63 |
| Log triglyceride |  |  |  |  |  |  |
| Overall | 15239 | 6641 |  |  |  |  |
| Birth | 2963 | 2963 | -0.71 (0.41) | -0.71 (0.15) | -0.001 | -0.52 to 0.51 |
| 0-9 years | 6808 | 5581 | -0.34 (0.53) | -0.35 (0.36) | 0.002 | -0.56 to 0.56 |
| 9-18 years | 8431 | 4595 | -0.14 (0.41) | -0.13 (0.22) | -0.002 | -0.56 to 0.55 |

SD, standard deviation

^a^ Insulin and triglyceride are natural log transformed. All values are in log form. Units for log insulin are in milliunits per litre. Units for log triglyceride are in millimoles per litre.

**Supplemental Table S7 Model details for HDL-c and non-HDL-c trajectories**

|  | No of contributing individuals | | Assessment of model fit | | | |
| --- | --- | --- | --- | --- | --- | --- |
|  | Total number of observations | Number of individuals with 1 measure | Mean observed HDL-c, mmol/l (SD) | Mean predicted HDL-c, mmol/l (SD) | Mean difference (observed – predicted), mmol/l | 95% level of agreement between observed and predicted, mmol/l |
| Overall | 15239 | 6641 |  |  |  |  |
| Birth | 2963 | 2963 | 0.52 (0.24) | 0.52 (0.11) | -0.000002 | -0.24 to 0.24 |
| 0-7 years | 6808 | 2965 | 1.09 (0.56) | 1.08 (0.52) | 0.01 | -0.24 to 0.25 |
| 7-18 years | 12274 | 5466 | 1.39 (0.32) | 1.39 (0.24) | -0.00002 | -0.25 to 0.25 |
| Overall | 15239 | 6641 |  |  |  |  |
| Birth | 2963 | 2963 | 1.22 (0.53) | 1.25 (0.25) | -0.02 | -0.59 to 0.54 |
| 0-9 years | 6808 | 5581 | 2.15 (1.00) | 2.10 (0.82) | 0.05 | -0.53 to 0.63 |
| 9-18 years | 8431 | 4595 | 2.63 (0.65) | 2.67 (0.52) | -0.04 | -0.59 to 0.51 |

HDL-c, high density lipoprotein cholesterol; Non-HDL-c, non-high density lipoprotein cholesterol; Mmol/l, millimoles per litre; SD, standard deviation.

**Supplemental Table S8 Number of participants with cardiometabolic health outcome measures at each time point**

|  | | Birth | | Age 1 | | Age 7 | | Age 9 | | Age 10 | | Age 11 | | | Age 12 | | Age 13 | | Age 15 | | Age 18 |
| --- | --- | --- | --- | --- | --- | --- | --- | --- | --- | --- | --- | --- | --- | --- | --- | --- | --- | --- | --- | --- | --- |
| BMI ^a^ | | |  | x | x | | x | | x | | x | | x | | | x | | x | | x | |
| Fat/lean mass | | |  |  |  | | 5,134 | |  | | 4,958 | |  | | | 4,348 | | 3,738 | | 3,437 | |
| SBP/DBP/pulse rate | | |  |  | 5,672 | | 5,347 | | 5,084 | | 4,970 | | 4,729 | | |  | | 3,787 | | 3,311 | |
| Glucose | | |  |  | 3,907 | | 662 | |  | |  | |  | | |  | | 2,485 | | 2,336 | |
| Lipids ^b^ | | | 2,863 |  | 3,846 | | 3,617 | |  | |  | |  | | |  | | 2,485 | | 2,328 | |
| Insulin | | | 206 |  | 440 | |  | |  | |  | |  | | |  | | 417 | | 401 | |

DBP, diastolic blood pressure; SBP, systolic blood pressure.

^a^ Measures available at each of these approximate ages and at several ages in between but exact timing and number of BMI measures not shown as measures were available from questionnaires, routine child health records and research clinics at different mean ages from 1 to 18 years.

^b^ Lipids include triglyceride, high density lipoprotein cholesterol (HDL-c) and non-HDL-c.

**Supplemental Table S9 Characteristics at birth of the mothers of children included in models compared with those excluded due to missing exposure, outcome or confounder data**

|  | **Participants included**  **n= 8,606** | **Participants excluded**  **n= 5,261 – 13,341 ^a^** |
| --- | --- | --- |
|  | **n (%)** | **n (%)** |
| **Household social class** |  |  |
| Professional | 1190(13.8) | 346(11.7) |
| Managerial & Technical | 3673(42.7) | 1148(38.9) |
| Non-Manual | 2254(26.2) | 690(23.4) |
| Manual | 1037(12.0) | 525(17.8) |
| Part Skilled & Unskilled | 452(5.3) | 240(8.1) |
| **Maternal education** |  |  |
| Less than O level | 2124(24.7) | 1624(42.1) |
| O level | 3167(36.8) | 1151(29.8) |
| A level | 2093(24.3) | 700(18.1) |
| Degree or above | 1222(14.2) | 382(9.9) |
| **Partners highest educational qualification** |  |  |
| Less than O level | 2488(29.8) | 1655(45.4) |
| O level | 1872(22.4) | 679(18.6) |
| A level | 2337(28.0) | 778(21.4) |
| Degree or Above | 1643(19.7) | 530(14.6) |
| **Maternal smoking during pregnancy** |  |  |
| No | 6781(78.8) | 3171(67.7) |
| Yes | 1825(21.2) | 1516(32.3) |
| **Parity** |  |  |
| 0 | 3893(45.2) | 1947(43.7) |
| 1 | 3159(36.7) | 1413(31.7) |
| 2 | 1554(18.1) | 1094(24.6) |
|  | **Mean (SD)** | **Mean (SD)** |
| Child gestational age at birth | 39.5(1.8) | 37(8.1) |
| Birthweight (g) | 3426(533) | 3310(643) |
| Maternal BMI (kg/m^2^) | 23.0(3.8) | 23(4.0) |
| Maternal age (years) | 28.7(4.7) | 27(5.2) |

SD, standard deviation

^a^ Denominators for excluded participants in this table vary due to different rates of missing data for characteristics shown.

**Supplemental Table S10 Mean trajectories of BMI estimated from multilevel models, by maternal anxiety during pregnancy**

|  | **Mean trajectory (95% CI) in males**  **(no anxiety during pregnancy) log (kg/m**^2^ **) (reference)** ^a^ | **Mean trajectory (95% CI) in females**  **(no anxiety during pregnancy) log (kg/m**^2^ **) (reference)** ^a^ | **Mean difference in trajectory (95% CI) comparing anxiety at 18 weeks only with the reference trajectory (% difference in kg/m**^2^**)** ^b^ | **Mean difference in trajectory (95% CI) anxiety at 32 weeks only with the reference trajectory (% difference in kg/m**^2^**)** ^b^ | **Mean difference in trajectory (95% CI) comparing anxiety at 18 and 32 weeks with the reference trajectory (% difference in kg/m**^2^**)** ^b^ |
| --- | --- | --- | --- | --- | --- |
| **Unadjusted** |  |  |  |  |  |
| **BMI** |  |  |  |  |  |
| Age 1yr | 2.90 (2.89,2.90) | 2.89 (2.88,2.89) | 0.42 (-1.00,1.84) | 0.13 (-1.15,1.40) | 0.3 (-0.85,1.53) |
| Age 3yr | 2.77 (2.76,2.77) | 2.75 (2.75,2.75) | 0.65 (-0.08,1.38) | 0.11 (-0.56,0.77) | 0.0 (-0.60,0.60) |
| Age 7yr | 2.79 (2.78,2.79) | 2.80 (2.79,2.80) | 0.95 (-0.01,1.91) | 0.44 (-0.43,1.32) | 0.6 (-0.20,1.39) |
| Age 9yr | 2.82 (2.82,2.83) | 2.84 (2.84,2.85) | 1.06 (-0.11,2.24) | 0.64 (-0.43,1.71) | 0.9 (-0.06,1.89) |
| Age 11yr | 2.87 (2.87,2.88) | 2.90 (2.90,2.91) | 1.16 (-0.18,2.50) | 0.83 (-0.39,2.06) | 1.2 (0.08,2.32) |
| Age 13yr | 2.94 (2.93,2.95) | 2.97 (2.96,2.97) | 1.23 (-0.21,2.66) | 1.03 (-0.28,2.34) | 1.4 (0.24,2.65) |
| Age 15yr | 3.02 (3.01,3.02) | 3.04 (3.04,3.05) | 1.27 (-0.23,2.76) | 1.22 (-0.16,2.60) | 1.6 (0.37,2.90) |
| Age 18yr | 3.16 (3.15,3.16) | 3.17 (3.16,3.18) | 1.27 (-0.51,3.05) | 1.50 (-0.16,3.17) | 1.8 (0.29,3.33) |
| **Adjusted** |  |  |  |  |  |
| **BMI** |  |  |  |  |  |
| Age 1yr | 2.88 (2.87,2.90) | 2.87 (2.86,2.89) | 0.36 (-1.06,1.79) | 0.18 (-1.11,1.47) | 0.3 (-0.91,1.51) |
| Age 3yr | 2.77 (2.76,2.77) | 2.75 (2.74,2.76) | 0.40 (-0.32,1.13) | 0.02 (-0.65,0.68) | -0.2 (-0.76,0.44) |
| Age 7yr | 2.78 (2.78,2.79) | 2.80 (2.79,2.81) | 0.43 (-0.49,1.34) | 0.27 (-0.57,1.11) | 0.4 (-0.36,1.17) |
| Age 9yr | 2.82 (2.81,2.83) | 2.84 (2.83,2.86) | 0.41 (-0.70,1.53) | 0.39 (-0.63,1.41) | 0.7 (-0.28,1.60) |
| Age 11yr | 2.87 (2.86,2.89) | 2.90 (2.89,2.91) | 0.38 (-0.88,1.64) | 0.49 (-0.66,1.65) | 0.8 (-0.22,1.91) |
| Age 13yr | 2.94 (2.92,2.95) | 2.97 (2.95,2.98) | 0.33 (-1.00,1.67) | 0.57 (-0.66,1.80) | 0.9 (-0.20,2.07) |
| Age 15yr | 3.02 (3.00,3.03) | 3.04 (3.03,3.06) | 0.26 (-1.13,1.65) | 0.61 (-0.67,1.90) | 0.9 (-0.27,2.10) |
| Age 18yr | 3.16 (3.14,3.18) | 3.17 (3.15,3.19) | 0.12 (-1.55,1.79) | 0.61 (-0.96,2.19) | 0.7 (-0.76,2.13) |

CI, confidence interval; yr, year.

^a^ BMI is presented in the natural log and values represent the mean predicted natural log of BMI at each age shown.

^b^ Differences at each age are back transformed from the log scale and are interpreted as the percentage difference in the mean level in original units at each age comparing each category with the reference trajectory.

**Supplemental Table S11 Mean trajectories of fat mass estimated from multilevel models, by maternal anxiety during pregnancy**

|  | **Mean trajectory (95% CI) in males**  **(no anxiety during pregnancy) log (kg or kg/yr) (reference)** ^a^ | **Mean trajectory (95% CI)**  **in females**  **(no anxiety during pregnancy) log (kg or kg/yr) (reference)** ^a^ | **Mean difference in trajectory (95% CI) comparing anxiety at 18 weeks only with the reference trajectory (% or %/yr)** ^b^ | **Mean difference in trajectory (95% CI) comparing anxiety at 32 weeks only with the reference trajectory (% or %/yr)** ^b^ | **Mean difference in trajectory (95% CI) comparing anxiety at 18 and 32 weeks with the reference trajectory (% or %/yr)** ^b^ |
| --- | --- | --- | --- | --- | --- |
| **Unadjusted** |  |  |  |  |  |
| Age 9yr (kg) or (%)  Change 9-13yr (kg/yr) or (%/yr) Change 13-15yr (kg/yr) or (%/yr) Change 15-18yr (kg/yr) or (%/yr) Age 18yr (kg) or (%) | 1.79 (1.76,1.82)  0.11 (0.11,0.12)  -0.06 (-0.07,-0.05)  0.10 (0.09,0.11)  2.43 (2.41,2.46) | 2.63 (2.11,3.15)  0.16 (0.15,0.17)  0.10 (0.09,0.11)  0.06 (0.05,0.06)  3.64 (3.09,4.18) | 3.62 (-2.42,9.67)  0.04 (-1.18,1.26)  1.41 (-0.84,3.66)  -1.41 (-3.04,0.21)  2.28 (-3.83,8.40) | -1.46 (-6.78,3.87)  1.00 (-0.15,2.15)  0.21 (-1.86,2.28)  -0.93 (-2.51,0.64)  0.11 (-5.56,5.79) | 0.94 (-4.08,5.95)  1.52 (0.44,2.60)  -1.62 (-3.49,0.26)  0.81 (-0.65,2.27)  6.33 (0.86,11.79) |
| **Adjusted** |  |  |  |  |  |
| Age 9yr (kg) or (%)  Change 9-13yr (kg/yr) or (%/yr)  Change 13-15yr (kg/yr) or (%/yr)  Change 15-18yr (kg/yr) or (%/yr)  Age 18yr (kg) or (%) | 1.81 (1.74,1.87)  0.11 (0.09,0.12)  -0.07 (-0.09,-0.04)  0.11 (0.09,0.12)  2.42 (2.35,2.48) | 2.66 (2.13,3.18)  0.15 (0.14,0.17)  0.09 (0.07,0.12)  0.06 (0.05,0.08)  3.63 (3.08,4.18) | 1.56 (-4.21,7.32)  -0.13 (-1.34,1.08)  1.56 (-0.70,3.82)  -1.47 (-3.10,0.16)  -0.32 (-6.10,5.45) | -2.02 (-7.17,3.14)  0.74 (-0.40,1.89)  0.21 (-1.87,2.28)  -0.93 (-2.51,0.64)  -1.46 (-6.88,3.97) | 1.00 (-3.92,5.91)  1.19 (0.12,2.27)  -1.75 (-3.64,0.14)  0.82 (-0.66,2.29)  4.77 (-0.51,10.05) |

CI, confidence interval; kg/yr, kilograms per year; %/yr, percentage per year.

^a^ Fat mass was transformed using the natural log. All predicted mean values (kg) and rates of change per year (kg/yr) for the reference categories are on the log scale

^b^ The difference between groups is back transformed from the log scale for ease of interpretation and is interpreted as the percentage difference in the mean level in original units comparing each category with the reference or percentage difference in change in original units per year (%/yr) comparing each category with the reference.

**Supplemental Table S12 Mean trajectories of blood pressure and pulse rate estimated from multilevel models, by maternal anxiety during pregnancy**

|  | Mean trajectory (95% CI) in males  (no anxiety during pregnancy) (reference) | Mean trajectory (95% CI) in females  (no anxiety during pregnancy) (reference) | Mean difference in trajectory (95% CI) comparing anxiety at 18 weeks only with the reference trajectory | | Mean difference in trajectory (95% CI) comparing anxiety at 32 weeks only with the reference trajectory | Mean difference in trajectory (95% CI) comparing anxiety at 18 and 32 weeks with the reference trajectory |
| --- | --- | --- | --- | --- | --- | --- |
| SBP |  |  |  | |  |  |
| Unadjusted |  |  |  |  | | 0.37 (-0.41,1.15)  -0.07 (-0.25,0.12)  -0.04 (-0.36,0.28)  0.42 (-0.30,1.15)  0.74 (-0.27,1.75) |
| Age 7yr (mmHg) | 97.76 (97.39,98.12)  1.65 (1.56,1.74)  5.86 (5.70,6.01)  -4.06 (-4.40,-3.71)  121.32 (120.84,121.80) | 97.86 (97.48,98.23) | 0.67 (-0.26,1.61)  0.08 (-0.14,0.31)  0.02 (-0.35,0.40)  0.13 (-0.71,0.96)  1.44 (0.27,2.61) | 0.34 (-0.52,1.20)  -0.01 (-0.22,0.19)  0.02 (-0.33,0.38)  -0.04 (-0.83,0.74)  0.28 (-0.82,1.38) | |  |
| Change 7-12yr (mmHg/yr) |  | 1.87 (1.78,1.95) |  |  |  |  |
| Change 12-16yr (mmHg/yr) |  | 3.85 (3.70,4.00) |  |  |  |  |
| Change 16-18yr (mmHg/yr) |  | -5.94 (-6.27,-5.61) |  |  |  |  |
| Age 18yr (mmHg) |  | 110.73 (110.27,111.18) |  |  |  |  |
| Adjusted |  |  |  |  | |  |
| Age 7yr (mmHg) | 98.91 (97.92,99.90)  1.47 (1.24,1.71)  5.97 (5.57,6.37)  -4.24 (-5.16,-3.32)  121.67 (120.40,122.95) | 99.04 (98.04,100.04) | 0.53 (-0.40,1.47)  0.07 (-0.15,0.29)  0.04 (-0.33,0.42)  0.04 (-0.80,0.88)  1.16 (-0.02,2.33) | 0.16 (-0.70,1.02)  -0.01 (-0.22,0.19)  0.06 (-0.30,0.41)  -0.16 (-0.95,0.63)  0.01 (-1.10,1.11) | | 0.23 (-0.55,1.01)  -0.06 (-0.25,0.13)  -0.0001 (-0.32,0.32)  0.22 (-0.52,0.95)  0.38 (-0.64,1.40) |
| Change 7-12yr (mmHg/yr) |  | 1.69 (1.45,1.93) |  |  |  |  |
| Change 12-16yr (mmHg/yr) |  | 3.97 (3.56,4.37) |  |  |  |  |
| Change 16-18yr (mmHg/yr) |  | -6.15 (-7.07,-5.23) |  |  |  |  |
| Age 18yr (mmHg) |  | 111.07 (109.79,112.35) |  |  |  |  |
| DBP |  |  |  |  | |  |
| Unadjusted |  |  |  |  | |  |
| Age 7yr (mmHg) | 56.06 (55.80,56.32)  0.14 (0.07,0.21)  2.87 (2.74,3.00)  -2.59 (-2.88,-2.30)  63.05 (62.70,63.40) | 56.91 (56.64,57.18) | 0.74 (0.06,1.41)  -0.02 (-0.19,0.16)  -0.06 (-0.38,0.26)  0.27 (-0.43,0.98)  0.97 (0.13,1.81) | 0.15 (-0.47,0.78)  0.05 (-0.11,0.21)  -0.03 (-0.33,0.27)  0.27 (-0.40,0.94)  0.84 (0.05,1.64) | | 0.40 (-0.17,0.96)  -0.02 (-0.17,0.12)  -0.17 (-0.45,0.10)  0.46 (-0.14,1.07)  0.52 (-0.20,1.24) |
| Change 7-12yr (mmHg/yr) |  | 0.12 (0.05,0.19) |  |  |  |  |
| Change 12-16yr (mmHg/yr) |  | 2.34 (2.21,2.47) |  |  |  |  |
| Change 16-18yr (mmHg/yr) |  | -1.17 (-1.45,-0.89) |  |  |  |  |
| Age 18yr (mmHg) |  | 63.65 (62.85,64.46) |  |  |  |  |
| Adjusted |  |  |  |  | |  |
| Age 7yr (mmHg) | 56.79 (56.08,57.51)  0.05 (-0.13,0.23)  2.95 (2.60,3.30)  -2.73 (-3.52,-1.95)  63.39 (62.46,64.31) | 57.65 (56.93,58.38) | 0.70 (0.02,1.38)  -0.02 (-0.19,0.15)  -0.02 (-0.34,0.30)  0.19 (-0.52,0.89)  0.89 (0.05,1.73) | | 0.08 (-0.55,0.70)  0.05 (-0.11,0.21)  0.03 (-0.27,0.33)  0.11 (-0.56,0.78)  0.67 (-0.13,1.46) | 0.33 (-0.24,0.90)  -0.02 (-0.16,0.13)  -0.09 (-0.37,0.19)  0.22 (-0.40,0.84)  0.31 (-0.42,1.04) |
| Change 7-12yr (mmHg/yr) |  | 0.03 (-0.15,0.22) |  |  |  |  |
| Change 12-16yr (mmHg/yr) |  | 2.43 (2.08,2.78) |  |  |  |  |
| Change 16-18yr (mmHg/yr) |  | -1.34 (-2.12,-0.55) |  |  |  |  |
| Age 18yr (mmHg) |  | 64.03 (62.85,65.21) |  |  |  |  |
| Pulse rate |  |  |  | |  |  |
| Unadjusted |  |  |  | |  |  |
| Age 7yr (bpm) | 82.33 (81.90,82.75)  -1.89 (-1.99,-1.79)  -0.71 (-0.87,-0.55)  -4.21 (-4.57,-3.85)  61.62 (61.10,62.15) | 85.59 (85.16,86.03) | 0.62 (-0.47,1.71)  0.01 (-0.26,0.27)  -0.09 (-0.48,0.30)  0.62 (-0.24,1.49)  1.53 (0.26,2.80) | | 1.18 (0.17,2.19)  -0.30 (-0.54,-0.06)  0.24 (-0.13,0.61)  -0.03 (-0.84,0.78)  0.57 (-0.62,1.77) | 0.68 (-0.23,1.59)  -0.08 (-0.30,0.14)  0.17 (-0.16,0.51)  -0.04 (-0.78,0.70)  0.92 (-0.17,2.01) |
| Change 7-12yr (bpm/yr) |  | -1.73 (-1.83,-1.63) |  |  |  |  |
| Change 12-16yr (bpm/yr) |  | -0.25 (-0.41,-0.10) |  |  |  |  |
| Change 16-18yr (bpm/yr) |  | -4.83 (-5.17,-4.49) |  |  |  |  |
| Age 18yr (bpm) |  | 66.27 (65.77,66.76) |  |  |  |  |
| Adjusted |  |  |  | |  |  |
| Age 7yr (bpm) | 82.57 (81.42,83.73)  -1.90 (-2.18,-1.62)  -1.05 (-1.47,-0.63)  -3.67 (-4.62,-2.73)  61.52 (60.13,62.91) | 85.80 (84.62,86.97) | 0.62 (-0.47,1.71)  0.01 (-0.25,0.27)  -0.09 (-0.48,0.30)  0.58 (-0.28,1.45)  1.48 (0.21,2.75) | | 1.13 (0.12,2.14)  -0.29 (-0.53,-0.05)  0.22 (-0.15,0.58)  -0.09 (-0.90,0.73)  0.37 (-0.83,1.56) | 0.68 (-0.24,1.60)  -0.06 (-0.29,0.16)  0.14 (-0.20,0.48)  -0.14 (-0.89,0.61)  0.65 (-0.45,1.75) |
| Change 7-12yr (bpm/yr) |  | -1.73 (-2.01,-1.45) |  |  |  |  |
| Change 12-16yr (bpm/yr) |  | -0.60 (-1.03,-0.17) |  |  |  |  |
| Change 16-18yr (bpm/yr) |  | -4.34 (-5.29,-3.39) |  |  |  |  |
| Age 18yr (bpm) |  | 66.06 (64.67,67.45) |  |  |  |  |

bpm, beats per minute; bpm/yr, beats per minute per year; CI, confidence interval; mmHg, millimetres of mercury; mmHg/yr, millimetres of mercury per year.

**Supplemental Table S13 Mean trajectories of lean mass estimated from multilevel models, by maternal anxiety during pregnancy**

|  | **Mean trajectory (95% CI) in males**  **(no anxiety during pregnancy) (reference)** | **Mean trajectory (95% CI) in females**  **(no anxiety during pregnancy) (reference)** | **Mean difference in trajectory (95% CI) comparing anxiety at 18 weeks only with the reference trajectory** | **Mean difference in trajectory (95% CI) comparing anxiety at 32 weeks only with the reference trajectory** | **Mean difference in trajectory (95% CI) comparing anxiety at 18 and 32 weeks with the reference trajectory** |
| --- | --- | --- | --- | --- | --- |
| **Unadjusted** |  |  |  |  |  |
| Age 9yr (kg)  Change 9-13yr (kg/yr)  Change 13-15yr (kg/yr)  Change 15-18yr (kg/yr)  Age 18yr (kg) | 23.89 (23.76,24.03)  2.30 (2.24,2.36)  7.69 (7.59,7.79)  2.47 (2.40,2.55)  55.91 (55.67,56.15) | 20.78 (20.65,20.91)  3.22 (3.17,3.28)  1.58 (1.46,1.69)  0.38 (0.31,0.45)  37.96 (37.72,38.20) | -0.12 (-0.42,0.18)  0.11 (-0.02,0.24)  -0.33 (-0.57, -0.08)  -0.0004 (-0.18,0.18)  -0.32 (-0.91,0.27) | -0.28 (-0.56,0.002)  -0.10 (-0.22,0.02)  0.09 (-0.14,0.32)  0.04 (-0.13,0.21)  -0.39 (-0.95,0.17) | -0.13 (-0.38,0.13)  0.04 (-0.07,0.15)  0.003 (-0.21,0.21)  -0.03 (-0.18,0.13)  -0.04 (-0.54,0.47) |
| **Adjusted** |  |  |  |  |  |
| Age 9yr (kg) | 23.84 (23.52,24.17)  2.30 (2.16,2.44)  7.48 (7.22,7.74)  2.63 (2.44,2.82)  55.90 (55.27,56.54) | 20.76 (20.43,21.08) | -0.14 (-0.44,0.16)  0.09 (-0.04,0.22)  -0.32 (-0.57,-0.07)  0.01 (-0.17,0.19)  -0.38 (-0.96,0.21) | -0.26 (-0.53,0.02)  -0.10 (-0.22,0.01)  0.09 (-0.14,0.32)  0.05 (-0.12,0.22)  -0.34 (-0.90,0.22) | -0.05 (-0.31,0.20)  0.04 (-0.07,0.15)  0.01 (-0.21,0.22)  -0.02 (-0.17,0.14)  0.07 (-0.44,0.58) |
| Change 9-13yr (kg/yr) |  | 3.22 (3.08,3.36) |  |  |  |
| Change 13-15yr (kg/yr) |  | 1.36 (1.09,1.63) |  |  |  |
| Change 15-18yr (kg/yr) |  | 0.54 (0.35,0.73) |  |  |  |
| Age 18yr (kg) |  | 37.99 (37.35,38.63) |  |  |  |

kg/yr, kilograms per year.

**Supplemental Table S14 Mean trajectories of glucose estimated from multilevel models, by maternal anxiety during pregnancy**

|  | Mean trajectory  (95% CI) in males (no anxiety during pregnancy) (reference) | Mean trajectory (95% CI) in females  (no anxiety during pregnancy) (reference) | Mean difference in trajectory (95% CI) comparing anxiety at 18 weeks only with the reference trajectory | Mean difference in trajectory (95% CI) comparing anxiety at 32 weeks only with the reference trajectory | Mean difference in trajectory (95% CI) c comparing anxiety at 18 and 32 weeks with the reference trajectory |
| --- | --- | --- | --- | --- | --- |
| Unadjusted |  |  |  |  |  |
| Age 7yr (mmol/l) | 4.19 (4.17,4.21)  0.15 (0.14,0.15)  -0.08 (-0.09, -0.07)  5.11 (5.09,5.14) | 4.13 (4.10,4.15) | -0.06 (-0.12,0.005)  0.007 (-0.005,0.02)  -0.003 (-0.04,0.03)  -0.01 (-0.08,0.05) | -0.003 (-0.06,0.06)  0.001 (-0.01,0.01)  -0.02 (-0.05,0.01)  -0.04 (-0.11,0.02) | 0.03 (-0.02,0.08)  -0.004 (-0.01,0.01)  0.01 (-0.02,0.04)  0.03 (-0.02,0.09) |
| Change 7-15yr (mmol/l/yr) |  | 0.13 (0.13,0.14) |  |  |  |
| Change 15-18yr (mmol/l/yr) |  | -0.10 (-0.11, -0.09) |  |  |  |
| Age 18yr (mmol/l) |  | 4.89 (4.86,4.92) |  |  |  |
| Adjusted |  |  |  |  |  |
| Age 7yr (mmol/l) | 4.21 (4.14,4.27)  0.14 (0.13,0.16)  -0.08 (-0.11, -0.04)  5.13 (5.06,5.20) | 4.14 (4.07,4.21) | -0.06 (-0.13,0.003)  0.01 (-0.005,0.02)  -0.003 (-0.04,0.03)  -0.02 (-0.08,0.05) | -0.005 (-0.06,0.05)  0.001 (-0.01,0.01)  -0.02 (-0.05,0.02)  -0.04 (-0.11,0.02) | 0.04 (-0.02,0.09)  -0.004 (-0.01,0.01)  0.01 (-0.02,0.04)  0.04 (-0.02,0.09) |
| Change 7-15yr (mmol/l/yr) |  | 0.13 (0.12,0.14) |  |  |  |
| Change 15-18yr (mmol/l/yr) |  | -0.10 (-0.13, -0.06) |  |  |  |
| Age 18yr (mmol/l) |  | 4.91 (4.84,4.98) |  |  |  |

CI, confidence interval; mmol/l, millimole per litre; mmol/l/year, millimoles per litre per year.

**Supplemental Table S15 Mean trajectories of insulin and triglyceride estimated from multilevel models, by maternal anxiety during pregnancy**

|  | **Mean trajectory (95% CI) in males**  **(no anxiety during pregnancy) (reference)** ^a^ | **Mean trajectory (95% CI) in females**  **(no anxiety during pregnancy) (reference)** ^a^ | **Mean difference in trajectory (95% CI) comparing anxiety at 18 weeks only with the reference trajectory (% or %/yr)** ^b^ | **Mean difference in trajectory (95% CI) comparing anxiety at 32 weeks only with the reference trajectory (% or %/yr)** ^b^ | **Mean difference in trajectory (95% CI) comparing anxiety at 18 and 32 weeks with the reference trajectory (% or %/yr)** ^b^ |
| --- | --- | --- | --- | --- | --- |
| **Insulin** |  |  |  |  |  |
| **Unadjusted** |  |  |  |  |  |
| Birth (mu/l or %) | 0.99 (0.89,1.09) 0.04 (0.03,0.06)  0.14 (0.12,0.16)  -0.14 (-0.18,-0.10)  1.76 (1.68,1.85) | 1.08 (0.99,1.18)  0.05 (0.03,0.06)  0.14 (0.12,0.16)  -0.12 (-0.16, -0.08)  1.98 (1.89,2.06) | -7.29 (-31.57,16.99)  4.08 (-0.12,8.29)  -5.18 (-10.14, -0.22)  5.80 (-5.20,16.80)  14.38 (-11.51,40.28) | -13.73 (-38.87,11.41)  2.82 (-1.64,7.27)  1.94 (-3.48,7.35)  -8.82 (-18.80,1.16)  -5.79 (-29.01,17.42) | 23.37 (-3.86,50.60)  -2.58 (-5.91,0.75)  -0.96 (-5.57,3.64)  -2.06 (-11.52,7.41)  -13.54 (-31.07,3.99) |
| Change 0-9yr (mu/l/yr or %/yr) |  |  |  |  |  |
| Change 9-15yr (mu/l/yr or %/yr)  Change 15-18yr (mu/l/yr or %/yr) |  |  |  |  |  |
| Age 18yr (mu/l or %) |  |  |  |  |  |
| **Adjusted** |  |  |  |  |  |
| Birth (mu/l or %) | 1.05 (0.78,1.32)  0.01 (-0.03,0.06)  0.19 (0.13,0.24)  -0.16 (-0.27,-0.05)  1.82 (1.59,2.05) | 1.19 (0.91,1.46)  0.02 (-0.03,0.06)  0.19 (0.13,0.24)  -0.14 (-0.24,-0.03)  2.04 (1.81,2.27) | -7.71 (-31.62,16.20)  3.94 (-0.23,8.10)  -4.62 (-9.68,0.44)  4.58 (-6.45,15.62)  12.52 (-13.06,38.11) | -13.80 (-38.45,10.85)  2.63 (-1.76,7.02)  1.58 (-3.85,7.01)  -7.53 (-17.55,2.48)  -5.44 (-28.32,17.45) | 23.58 (-3.05,50.21)  -2.66 (-5.93,0.62)  -0.96 (-5.60,3.67)  -1.89 (-11.31,7.53)  -13.57 (-30.83,3.70) |
| Change 0-9yr (mu/l/yr or %/yr) |  |  |  |  |  |
| Change 9-15yr (mu/l/yr or %/yr)  Change 15-18yr (mu/l/yr or %/yr) |  |  |  |  |  |
| Age 18yr (mu/l or %) |  |  |  |  |  |
| **Triglyceride** |  |  |  |  |  |
| **Unadjusted** |  |  |  |  |  |
| Birth (mmol/l or %) | -0.71 (-0.73,-0.69)  0.08 (0.08,0.09)  -0.04 (-0.04,-0.04)  -0.33 (-0.35,-0.31) | -0.70 (-0.72,-0.68) | -0.25 (-5.56,5.06)  0.14 (-0.67,0.95)  0.36 (-0.37,1.09)  4.34 (-0.82,9.51) | -3.01 (-8.22,2.20)  0.19 (-0.60,0.98)  0.08 (-0.60,0.77)  -0.60 (-5.30,4.11) | 1.66 (-2.92,6.24)  -0.35 (-1.04,0.33)  0.14 (-0.49,0.78)  -0.26 (-4.66,4.13) |
| Change 0-9yr (mmol/l/yr or %/yr) |  | 0.09 (0.09,0.09) |  |  |  |
| Change 9-18yr (mmol/l/yr or %/yr) |  | -0.04 (-0.05,-0.04) |  |  |  |
| Age 18yr (mmol/l or %) |  | -0.30 (-0.32,-0.28) |  |  |  |
| **Adjusted** |  |  |  |  |  |
| Birth (mmol/l or %) | -0.66 (-0.72,-0.60)  0.08 (0.07,0.09)  -0.04 (-0.05,-0.04)  -0.32 (-0.37,-0.26) | -0.65 (-0.71,-0.59)  0.09 (0.08,0.10)  -0.05 (-0.05,-0.04)  -0.28 (-0.34,-0.23) | -1.50 (-6.63,3.63)  0.25 (-0.55,1.06)  0.28 (-0.45,1.00)  3.29 (-1.83,8.42) | -2.86 (-7.96,2.25)  0.16 (-0.62,0.95)  0.02 (-0.67,0.71)  -1.24 (-5.92,3.44) | 1.15 (-3.35,5.65)  -0.27 (-0.96,0.41)  -0.01 (-0.65,0.63)  -1.35 (-5.73,3.04) |
| Change 0-9yr (mmol/l/yr or %/yr) |  |  |  |  |  |
| Change 9-18yr (mmol/l/yr or %/yr) |  |  |  |  |  |
| Age 18yr (mmol/l or %) |  |  |  |  |  |

CI, confidence interval; mmol/l, millimole per litre; mmol/l/year, millimoles per litre per year; %/yr, percentage per year

^a^ Insulin and triglyceride were transformed using the natural log. All predicted mean values and rates of change per year for the reference categories are on the log scale.

^b^ The difference between groups is back transformed from the log scale for ease of interpretation and is interpreted as the percentage difference in the mean level in original units comparing each category with the reference or percentage difference in change in original units per year (%/yr) comparing each category with the reference.

**Supplemental Table S16 Mean trajectories of HDL-C and non-HDL-c estimated from multilevel models, by maternal anxiety during pregnancy**

|  | **Mean trajectory (95% CI) in males**  **(no anxiety during pregnancy) (reference)** | **Mean trajectory (95% CI) in females**  **(no anxiety during pregnancy) (reference)** | **Mean difference in trajectory (95% CI) comparing anxiety at 18 weeks only with the reference trajectory** | **Mean difference in trajectory (95% CI) comparing anxiety at 32 weeks only with the reference trajectory** | **Mean difference in trajectory (95% CI) c comparing anxiety at 18 and 32 weeks with the reference trajectory** |
| --- | --- | --- | --- | --- | --- |
| **HDL-c** |  |  |  |  |  |
| **Unadjusted** |  |  |  |  |  |
| Birth (mmol/l) | 0.50 (0.49,0.51)  0.15 (0.15,0.15)  -0.04 (-0.04,-0.03)  1.15 (1.14,1.17) | 0.55 (0.53,0.56) | 0.01 (-0.02,0.04)  -0.005 (-0.01,0.002)  0.0003 (-0.003,0.004)  -0.02 (-0.05,0.02) | 0.01 (-0.02,0.04)  0.0004 (-0.01,0.01)  -0.002 (-0.01,0.001)  -0.01 (-0.05,0.02) | -0.003 (-0.03,0.02)  -0.001 (-0.01,0.004)  -0.00003 (-0.003,0.003)  -0.01 (-0.04,0.02) |
| Change 0-7yr (mmol/l/yr) |  | 0.13 (0.13,0.13) |  |  |  |
| Change 7-18yr (mmol/l/yr) |  | -0.01 (-0.01,-0.01) |  |  |  |
| Age 18yr (mmol/l) |  | 1.33 (1.32,1.35) |  |  |  |
| **Adjusted** |  |  |  |  |  |
| Birth (mmol/l) | 0.49 (0.46,0.53)  0.15 (0.14,0.16)  -0.04 (-0.04,-0.03)  1.16 (1.13,1.20) | 0.54 (0.50,0.57) | 0.02 (-0.02,0.05)  -0.005 (-0.01,0.001)  0.001 (-0.003,0.004)  -0.01 (-0.05,0.03) | 0.01 (-0.02,0.04)  0.001 (-0.01,0.01)  -0.001 (-0.005,0.002)  -0.005 (-0.04,0.03) | 0.003 (-0.02,0.03)  -0.002 (-0.01,0.003)  0.001 (-0.002,0.004)  0.0005 (-0.03,0.03) |
| Change 0-7yr (mmol/l/yr) |  | 0.13 (0.13,0.14) |  |  |  |
| Change 7-18yr (mmol/l/yr) |  | -0.01 (-0.01,-0.01) |  |  |  |
| Age 18yr (mmol/l) |  | 1.35 (1.31,1.39) |  |  |  |
| **Non-HDL-c** |  |  |  |  |  |
| **Unadjusted** |  |  |  |  |  |
| Birth (mmol/l) | 1.23 (1.20,1.26)  0.19 (0.18,0.19)  -0.07 (-0.08,-0.07)  2.27 (2.23,2.30) | 1.30 (1.27,1.33) | -0.08 (-0.18,0.01)  0.017 (0.001,0.03)  0.004 (-0.01,0.02)  0.11 (-0.01,0.22) | 0.03 (-0.08,0.13)  -0.005 (-0.02,0.01)  0.01 (-0.01,0.02)  0.04 (-0.07,0.15) | -0.02 (-0.10,0.06)  -0.002 (-0.01,0.01)  0.01 (-0.01,0.02)  0.02 (-0.08,0.12) |
| Change 0-9yr (mmol/l/yr) |  | 0.21 (0.20,0.21) |  |  |  |
| Change 9-18yr (mmol/l/yr) |  | -0.07 (-0.08,-0.07) |  |  |  |
| Age 18yr (mmol/l) |  | 2.49 (2.45,2.52) |  |  |  |
| **Adjusted** |  |  |  |  |  |
| Birth (mmol/l) | 1.21 (1.13,1.29)  0.19 (0.18,0.20)  -0.07 (-0.08,-0.06)  2.27 (2.18,2.36) | 1.27 (1.19,1.35) | -0.09 (-0.19,0.01)  0.02 (0.002,0.03)  0.002 (-0.01,0.02)  0.09 (-0.02,0.20) | 0.03 (-0.08,0.13)  -0.004 (-0.02,0.01)  0.01 (-0.01,0.02)  0.04 (-0.07,0.15) | -0.02 (-0.10,0.06)  -0.003 (-0.02,0.01)  0.005 (-0.01,0.02)  0.001 (-0.10,0.10) |
| Change 0-9yr (mmol/l/yr) |  | 0.21 (0.20,0.22) |  |  |  |
| Change 9-18yr (mmol/l/yr) |  | -0.07 (-0.08,-0.06) |  |  |  |
| Age 18yr (mmol/l) |  | 2.49 (2.40,2.58) |  |  |  |

CI, confidence interval; mmol/l, millimole per litre; mmol/l, millimole per litre per year.

**Supplemental Table S17 Mean trajectories of BMI estimated from multilevel models, by maternal depression during pregnancy**

|  | **Mean trajectory (95% CI)**  **in males**  **(no depression during pregnancy) log (kg/m**^2^ **) (reference)** ^a^ | | **Mean trajectory (95% CI)**  **in females**  **(no depression during pregnancy) log (kg/m**^2^ **) (reference)** ^a^ | | **Mean difference in trajectory (95% CI) comparing depression at 18 weeks only with the reference trajectory (% difference in kg/m**^2^**)** ^b^ | **Mean difference in trajectory (95% CI) depression at 32 weeks only with the reference trajectory (% difference in kg/m**^2^**)** ^b^ | **Mean difference in trajectory (95% CI) comparing depression at 18 and 32 weeks with the reference trajectory (% difference in kg/m**^2^**)** ^b^ |
| --- | --- | --- | --- | --- | --- | --- | --- |
| **Unadjusted** | |  | |  |  |  |  |
| **BMI** | |  | |  |  |  |  |
| Age 1yr | | 2.90 (2.89,2.91) | | 2.89 (2.88,2.89) | 0.07 (-1.60,1.75) | -0.33 (-1.79,1.13) | 0.3 (-1.32,1.91) |
| Age 3yr | | 2.77 (2.76,2.77) | | 2.75 (2.75,2.76) | 0.13 (-0.72,0.98) | 0.39 (-0.36,1.15) | -0.5 (-1.34,0.26) |
| Age 7yr | | 2.79 (2.78,2.79) | | 2.80 (2.79,2.80) | 0.51 (-0.61,1.62) | 0.61 (-0.38,1.59) | 0.1 (-0.97,1.11) |
| Age 9yr | | 2.82 (2.82,2.83) | | 2.84 (2.84,2.85) | 0.70 (-0.67,2.07) | 0.71 (-0.50,1.92) | 0.5 (-0.77,1.80) |
| Age 11yr | | 2.88 (2.87,2.88) | | 2.90 (2.90,2.91) | 0.88 (-0.69,2.44) | 0.85 (-0.53,2.23) | 1.0 (-0.48,2.46) |
| Age 13yr | | 2.94 (2.94,2.95) | | 2.97 (2.96,2.98) | 1.04 (-0.64,2.72) | 1.02 (-0.46,2.50) | 1.5 (-0.10,3.07) |
| Age 15yr | | 3.02 (3.01,3.02) | | 3.04 (3.04,3.05) | 1.19 (-0.58,2.95) | 1.24 (-0.31,2.80) | 2.0 (0.31,3.66) |
| Age 18yr | | 3.16 (3.15,3.16) | | 3.17 (3.16,3.18) | 1.37 (-0.77,3.50) | 1.67 (-0.21,3.55) | 2.7 (0.71,4.78) |
| **Adjusted** | |  | |  |  |  |  |
| **BMI** | |  | |  |  |  |  |
| Age 1yr | | 2.88 (2.87,2.90) | | 2.87 (2.86,2.89) | 0.04 (-1.66,1.73) | -0.37 (-1.83,1.10) | 0.2 (-1.45,1.82) |
| Age 3yr | | 2.77 (2.76,2.78) | | 2.75 (2.74,2.76) | -0.19 (-1.04,0.66) | 0.24 (-0.52,0.99) | -0.8 (-1.63,-0.04) |
| Age 7yr | | 2.79 (2.78,2.80) | | 2.80 (2.79,2.81) | -0.15 (-1.22,0.92) | 0.29 (-0.66,1.23) | -0.5 (-1.49,0.52) |
| Age 9yr | | 2.82 (2.81,2.83) | | 2.84 (2.83,2.86) | -0.14 (-1.44,1.16) | 0.28 (-0.87,1.43) | -0.2 (-1.44,1.00) |
| Age 11yr | | 2.87 (2.86,2.89) | | 2.90 (2.89,2.92) | -0.15 (-1.62,1.32) | 0.27 (-1.03,1.57) | 0.0 (-1.35,1.43) |
| Age 13yr | | 2.94 (2.92,2.95) | | 2.97 (2.96,2.98) | -0.18 (-1.75,1.39) | 0.28 (-1.11,1.66) | 0.3 (-1.21,1.76) |
| Age 15yr | | 3.02 (3.00,3.03) | | 3.05 (3.03,3.06) | -0.24 (-1.88,1.40) | 0.30 (-1.16,1.75) | 0.5 (-1.09,2.03) |
| Age 18yr | | 3.16 (3.14,3.18) | | 3.17 (3.16,3.19) | -0.38 (-2.39,1.63) | 0.37 (-1.41,2.15) | 0.7 (-1.23,2.61) |

CI, confidence interval; yr, year.

^a^ BMI is presented in the natural log and values represent the mean predicted natural log of BMI at each age shown.

^b^ Differences at each age are back transformed from the log scale and are interpreted as the percentage difference in the mean level in original units at each age comparing each category with the reference trajectory.

**Supplemental Table S18 Mean trajectories of fat mass estimated from multilevel models, by maternal depression during pregnancy**

|  | **Mean trajectory (95% CI) in males**  **(no depression during pregnancy) log (kg or kg/yr) (reference)** ^a^ | **Mean trajectory (95% CI) in females**  **(no depression during pregnancy) log (kg or kg/yr) (reference)** ^a^ | **Mean difference in trajectory (95% CI) comparing depression at 18 weeks only with the reference trajectory (% or %/yr)** ^b^ | **Mean difference in trajectory (95% CI) depression at 32 weeks only with the reference trajectory (% or %/yr)** ^b^ | **Mean difference in trajectory (95% CI) comparing depression at 18 and 32 weeks with the reference trajectory (% or %/yr)** ^b^ |
| --- | --- | --- | --- | --- | --- |
| **Unadjusted** |  |  |  |  |  |
| Age 9yr (kg) or (%)  Change 9-13yr (kg/yr) or (%/yr)  Change 13-15yr (kg/yr) or (%/yr)  Change 15-18yr (kg/yr) or (%/yr)  Age 18yr (kg) or (%) | 1.79 (1.76,1.82)  0.12 (0.11,0.12)  -0.06 (-0.07,-0.05)  0.10 (0.09,0.10)  2.43 (2.41,2.46) | 2.64 (2.12,3.16)  0.16 (0.15,0.17)  0.10 (0.09,0.11)  0.06 (0.05,0.06)  3.65 (3.10,4.20) | 2.03 (-5.19,9.26)  0.90 (-0.60,2.41)  -2.44 (-5.07,0.20)  1.23 (-0.83,3.28)  4.43 (-3.14,12.01) | -3.60 (-9.47,2.28)  0.77 (-0.52,2.06)  0.22 (-2.08,2.52)  -0.28 (-2.03,1.48)  -0.98 (-7.26,5.30) | 1.61 (-5.11,8.33)  1.86 (0.40,3.32)  -0.61 (-3.24,2.03)  -0.44 (-2.43,1.54)  6.61 (-0.74,13.96) |
| **Adjusted** |  |  |  |  |  |
| Age 9yr (kg) or (%)  Change 9-13yr (kg/yr) or (%/yr)  Change 13-15yr (kg/yr) or (%/yr)  Change 15-18yr (kg/yr) or (%/yr)  Age 18yr (kg) or (%) | 1.81 (1.74,1.87)  0.11 (0.09,0.12)  -0.07 (-0.09,-0.04)  0.10 (0.09,0.12)  2.42 (2.36,2.48) | 2.67 (2.14,3.19)  0.15 (0.14,0.17)  0.09 (0.07,0.12)  0.06 (0.04,0.08)  3.64 (3.09,4.19) | -0.13 (-7.03,6.77)  0.56 (-0.94,2.06)  -2.41 (-5.05,0.24)  1.25 (-0.82,3.32)  0.96 (-6.17,8.09) | -3.49 (-9.23,2.25)  0.37 (-0.92,1.66)  -0.02 (-2.32,2.29)  -0.20 (-1.97,1.56)  -2.67 (-8.69,3.34) | -0.21 (-6.66,6.24)  1.33 (-0.12,2.79)  -0.78 (-3.42,1.87)  -0.41 (-2.41,1.59)  2.32 (-4.58,9.22) |

CI, confidence interval; kg/yr, kilograms per year; %/yr, percentage per year.

^a^ Fat mass was transformed using the natural log. All predicted mean values (kg) and rates of change per year (kg/yr) for the reference categories are on the log scale

^b^ The difference between groups is back transformed from the log scale for ease of interpretation and is interpreted as the percentage difference in the mean level in original units comparing each category with the reference or percentage difference in change in original units per year (%/yr) comparing each category with the reference.

**Supplemental Table S19 Mean trajectories of blood pressure and pulse rate estimated from multilevel models, by maternal depression during pregnancy**

|  | Mean trajectory (95% CI) in males  (no depression during pregnancy) (reference) | Mean trajectory (95% CI) in females  (no depression during pregnancy) (reference) | Mean difference in trajectory (95% CI) comparing depression at 18 weeks only with the reference trajectory | Mean difference in trajectory (95% CI) comparing depression at 32 weeks only with the reference trajectory | Mean difference in trajectory (95% CI) comparing depression at 18 and 32 weeks with the reference trajectory | |
| --- | --- | --- | --- | --- | --- | --- |
| SBP |  |  |  |  |  | |
| Unadjusted |  |  | -0.18 (-1.31,0.95)  0.17 (-0.10,0.44)  0.11 (-0.35,0.58)  0.48 (-0.55,1.52)  2.11 (0.67,3.55) | 0.33 (-0.64,1.30)  -0.13 (-0.36,0.10)  0.17 (-0.23,0.56)  -0.64 (-1.53,0.24)  -0.93 (-2.17,0.31) | | 0.30 (-0.75,1.35)  -0.14 (-0.39,0.11)  0.38 (-0.05,0.82)  -0.07 (-1.04,0.91)  1.00 (-0.36,2.36) |
| Age 7yr (mmHg) | 97.85 (97.49,98.20)  1.66 (1.57,1.74)  5.82 (5.67,5.97)  -3.98 (-4.32,-3.65)  121.44 (120.97,121.91) | 97.95 (97.59,98.31) |  |  |  |  |
| Change 7-12yr (mmHg/yr) |  | 1.87 (1.79,1.96) |  |  |  |  |
| Change 12-16yr (mmHg/yr) |  | 3.81 (3.67,3.96) |  |  |  |  |
| Change 16-18yr (mmHg/yr) |  | -5.86 (-6.18,-5.54) |  |  |  |  |
| Age 18yr (mmHg) |  | 110.85 (110.42,111.28) |  |  |  |  |
| Adjusted |  |  |  |  | |  |
| Age 7yr (mmHg) | 98.97 (97.99,99.96)  1.48 (1.24,1.71)  5.95 (5.55,6.35)  -4.20 (-5.11,-3.29)  121.76 (120.49,123.03) | 99.11 (98.11,100.10)  1.70 (1.46,1.94)  3.94 (3.53,4.35)  -6.11 (-7.02,-5.19)  111.15 (109.88,112.42) | -0.46 (-1.59,0.66)  0.18 (-0.09,0.45)  0.18 (-0.28,0.65)  0.23 (-0.80,1.27)  1.62 (0.17,3.07) | 0.16 (-0.80,1.13)  -0.12 (-0.35,0.11)  0.23 (-0.17,0.63)  -0.88 (-1.77,0.01)  -1.29 (-2.54,-0.05) | | 0.02 (-1.03,1.07)  -0.14 (-0.39,0.11)  0.44 (-0.002,0.87)  -0.28 (-1.26,0.70)  0.50 (-0.87,1.87) |
| Change 7-12yr (mmHg/yr) |  |  |  |  |  |  |
| Change 12-16yr (mmHg/yr) |  |  |  |  |  |  |
| Change 16-18yr (mmHg/yr) |  |  |  |  |  |  |
| Age 18yr (mmHg) |  |  |  |  |  |  |
| DBP |  |  |  |  | |  |
| Unadjusted |  |  |  |  | |  |
| Age 7yr (mmHg) | 56.11 (55.86,56.36)  0.14 (0.07,0.21)  2.85 (2.73,2.98)  -2.55 (-2.83,-2.26)  63.14 (62.80,63.48) | 56.96 (56.70,57.22) | 0.24 (-0.58,1.06)  0.04 (-0.17,0.25)  0.06 (-0.34,0.45)  0.17 (-0.69,1.03)  0.99 (-0.03,2.02) | 0.44 (-0.26,1.14)  0.03 (-0.15,0.21)  -0.12 (-0.46,0.22)  0.02 (-0.72,0.77)  0.18 (-0.72,1.07) | | 0.31 (-0.45,1.07)  -0.07 (-0.27,0.12)  -0.10 (-0.47,0.27)  0.70 (-0.12,1.51)  0.95 (-0.02,1.91) |
| Change 7-12yr (mmHg/yr) |  | 0.12 (0.05,0.19) |  |  |  |  |
| Change 12-16yr (mmHg/yr) |  | 2.33 (2.20,2.45) |  |  |  |  |
| Change 16-18yr (mmHg/yr) |  | -1.12 (-1.39,-0.85) |  |  |  |  |
| Age 18yr (mmHg) |  | 63.76 (62.96,64.56) |  |  |  |  |
| Adjusted |  |  |  |  | |  |
| Age 7yr (mmHg) | 56.83 (56.11,57.54)  0.05 (-0.13,0.23)  2.94 (2.60,3.29)  -2.70 (-3.48,-1.92)  63.46 (62.54,64.39) | 57.69 (56.97,58.41) | 0.14 (-0.68,0.96)  0.03 (-0.18,0.24)  0.15 (-0.24,0.55)  -0.09 (-0.96,0.78)  0.73 (-0.30,1.77) | 0.37 (-0.33,1.07)  0.03 (-0.14,0.21)  -0.02 (-0.36,0.32)  -0.27 (-1.02,0.48)  -0.07 (-0.96,0.83) | 0.20 (-0.56,0.96)  -0.08 (-0.27,0.12)  0.02 (-0.36,0.40)  0.34 (-0.48,1.17)  0.59 (-0.38,1.56) | |
| Change 7-12yr (mmHg/yr) |  | 0.04 (-0.15,0.22) |  |  |  |  |
| Change 12-16yr (mmHg/yr) |  | 2.42 (2.07,2.77) |  |  |  |  |
| Change 16-18yr (mmHg/yr) |  | -1.31 (-2.08,-0.53) |  |  |  |  |
| Age 18yr (mmHg) |  | 64.13 (62.95,65.30) |  |  |  |  |
| Pulse rate |  |  |  |  |  | |
| Unadjusted |  |  |  |  |  | |
| Age 7yr (bpm) | 82.49 (82.08,82.90)  -1.92 (-2.02,-1.82)  -0.73 (-0.88,-0.57)  -4.08 (-4.43,-3.73)  61.82 (61.31,62.33) | 85.76 (85.34,86.18) | 0.36 (-0.96,1.68)  -0.14 (-0.46,0.18)  0.43 (-0.05,0.92)  -0.65 (-1.72,0.41)  0.07 (-1.49,1.63) | 0.51 (-0.62,1.64)  0.03 (-0.24,0.30)  0.24 (-0.17,0.65)  -0.83 (-1.74,0.08)  -0.04 (-1.38,1.31) | 0.07 (-1.16,1.29)  0.08 (-0.22,0.38)  0.29 (-0.17,0.74)  -0.19 (-1.19,0.81)  1.22 (-0.25,2.68) | |
| Change 7-12yr (bpm/yr) |  | -1.76 (-1.86,-1.66) |  |  |  |  |
| Change 12-16yr (bpm/yr) |  | -0.28 (-0.43,-0.12) |  |  |  |  |
| Change 16-18yr (bpm/yr) |  | -4.69 (-5.02,-4.36) |  |  |  |  |
| Age 18yr (bpm) |  | 66.46 (65.99,66.93) |  |  |  |  |
| Adjusted |  |  |  |  |  | |
| Age 7yr (bpm) | 82.71 (81.56,83.86)  -1.93 (-2.21,-1.66)  -1.05 (-1.47,-0.63)  -3.61 (-4.56,-2.67)  61.63 (60.25,63.01) | 85.94 (84.77,87.11) | 0.24 (-1.09,1.56)  -0.11 (-0.43,0.21)  0.42 (-0.07,0.91)  -0.74 (-1.80,0.33)  -0.12 (-1.68,1.44) | 0.51 (-0.62,1.64)  0.04 (-0.23,0.31)  0.21 (-0.21,0.62)  -0.91 (-1.82,0.01)  -0.26 (-1.61,1.09) | 0.05 (-1.19,1.28)  0.08 (-0.22,0.38)  0.26 (-0.19,0.72)  -0.29 (-1.29,0.71)  0.93 (-0.54,2.40) | |
| Change 7-12yr (bpm/yr) |  | -1.76 (-2.04,-1.48) |  |  |  |  |
| Change 12-16yr (bpm/yr) |  | -0.60 (-1.03,-0.17) |  |  |  |  |
| Change 16-18yr (bpm/yr) |  | -4.27 (-5.21,-3.33) |  |  |  |  |
| Age 18yr (bpm) |  | 66.17 (64.79,67.55) |  |  |  |  |

bpm, beats per minute; bpm/yr, beats per minute per year; CI, confidence interval; mmHg, millimetres of mercury; mmHg/yr, millimetres of mercury per year.

**Supplemental Table S20 Mean trajectories of lean mass estimated from multilevel models, by maternal depression during pregnancy**

|  | **Mean trajectory (95% CI) in males**  **(no depression during pregnancy) (reference)** | **Mean trajectory (95% CI) in females**  **(no depression during pregnancy) (reference)** | **Mean difference in trajectory (95% CI) comparing depression at 18 weeks only with the reference trajectory** | **Mean difference in trajectory (95% CI) comparing depression at 32 weeks only with the reference trajectory** | **Mean difference in trajectory (95% CI) comparing depression at 18 and 32 weeks with the reference trajectory** |
| --- | --- | --- | --- | --- | --- |
| **Unadjusted** |  |  |  |  |  |
| Age 9yr (kg) | 23.89 (23.76,24.02)  2.31 (2.25,2.37)  7.68 (7.59,7.78)  2.47 (2.40,2.55)  55.91 (55.68,56.14) | 20.77 (20.65,20.90) | -0.10 (-0.47,0.26)  0.03 (-0.13,0.18)  -0.05 (-0.35,0.25)  -0.01 (-0.23,0.21)  -0.13 (-0.85,0.58) | -0.39 (-0.70,-0.07)  -0.01 (-0.14,0.13)  -0.02 (-0.28,0.23)  0.002 (-0.19,0.19)  -0.46 (-1.09,0.17) | -0.18 (-0.53,0.17)  -0.03 (-0.17,0.12)  -0.04 (-0.33,0.26)  0.0005 (-0.21,0.22)  -0.35 (-1.03,0.33) |
| Change 9-13yr (kg/yr) |  | 3.23 (3.17,3.28) |  |  |  |
| Change 13-15yr (kg/yr) |  | 1.57 (1.46,1.68) |  |  |  |
| Change 15-18yr (kg/yr) |  | 0.38 (0.31,0.44) |  |  |  |
| Age 18yr (kg) |  | 37.95 (37.72,38.18) |  |  |  |
| **Adjusted** |  |  |  | -0.33 (-0.65,-0.02)  -0.01 (-0.14,0.13)  -0.03 (-0.29,0.23)  0.01 (-0.18,0.20)  -0.40 (-1.02,0.23) |  |
| Age 9yr (kg) | 23.83 (23.51,24.16)  2.30 (2.16,2.44)  7.47 (7.21,7.73)  2.64 (2.45,2.83)  55.90 (55.27,56.53) | 20.74 (20.42,21.07) | -0.06 (-0.43,0.30)  0.0001 (-0.16,0.16)  -0.03 (-0.33,0.27)  0.002 (-0.22,0.22)  -0.11 (-0.83,0.60) |  | -0.16 (-0.51,0.18)  -0.04 (-0.18,0.11)  -0.02 (-0.32,0.27)  0.01 (-0.21,0.22)  -0.33 (-1.02,0.35) |
| Change 9-13yr (kg/yr) |  | 3.22 (3.08,3.36) |  |  |  |
| Change 13-15yr (kg/yr) |  | 1.35 (1.08,1.62) |  |  |  |
| Change 15-18yr (kg/yr) |  | 0.55 (0.36,0.74) |  |  |  |
| Age 18yr (kg) |  | 37.98 (37.34,38.61) |  |  |  |

kg/yr, kilograms per year.

**Supplemental Table S21 Mean trajectories of glucose estimated from multilevel models, by maternal depression during pregnancy**

|  | Mean trajectory (95% CI) in males  (no depression during pregnancy) (reference) | Mean trajectory (95% CI) in females  (no depression during pregnancy) (reference) | Mean difference in trajectory (95% CI) comparing depression at 18 weeks only with the reference trajectory | Mean difference in trajectory (95% CI) comparing depression at 32 weeks only with the reference trajectory | Mean difference in trajectory (95% CI) c comparing depression at 18 and 32 weeks with the reference trajectory |
| --- | --- | --- | --- | --- | --- |
| Unadjusted |  |  |  |  |  |
| Age 7yr (mmol/l) | 4.20 (4.17,4.22)  0.14 (0.14,0.15)  -0.08 (-0.09,-0.07)  5.11 (5.09,5.14) | 4.13 (4.11,4.15) | -0.04 (-0.12,0.04)  0.001 (-0.01,0.01)  0.01 (-0.03,0.05)  -0.01 (-0.08,0.07) | -0.05 (-0.11,0.02)  0.002 (-0.01,0.01)  - 0.003 (-0.04,0.03)  -0.03 (-0.10,0.04) | -0.04 (-0.11,0.04)  0.01 (-0.002,0.03)  -0.01 (-0.05,0.03)  0.02 (-0.06,0.10) |
| Change 7-15yr (mmol/l/yr) |  | 0.13 (0.13,0.14) |  |  |  |
| Change 15-18yr (mmol/l/yr) |  | -0.10 (-0.11,-0.09) |  |  |  |
| Age 18yr (mmol/l) |  | 4.89 (4.87,4.92) |  |  |  |
| Adjusted |  |  |  |  |  |
| Age 7yr (mmol/l) | 4.21 (4.14,4.28)  0.14 (0.13,0.16)  -0.08 (-0.11,-0.04)  5.13 (5.06,5.20) | 4.14 (4.08,4.21) | -0.04 (-0.12,0.03)  0.001 (-0.01,0.01)  0.01 (-0.03,0.05)  -0.01 (-0.09,0.06) | -0.04 (-0.11,0.02)  0.002 (-0.01,0.01)  -0.001 (-0.04,0.03)  -0.03 (-0.10,0.04) | -0.03 (-0.10,0.04)  0.01 (-0.002,0.03)  -0.01 (-0.05,0.03)  0.02 (-0.06,0.10) |
| Change 7-15yr (mmol/l/yr) |  | 0.13 (0.12,0.14) |  |  |  |
| Change 15-18yr (mmol/l/yr) |  | -0.10 (-0.13,-0.06) |  |  |  |
| Age 18yr (mmol/l) |  | 4.91 (4.84,4.98) |  |  |  |

CI, confidence interval; mmol/l, millimole per litre; mmol/l/year, millimoles per litre per year.

**Supplemental Table S22 Mean trajectories of insulin and triglyceride estimated from multilevel models, by maternal depression during pregnancy**

|  | **Mean trajectory (95% CI) in males**  **(no depression during pregnancy) (reference)** ^a^ | **Mean trajectory (95% CI) in females**  **(no depression during pregnancy) (reference)** ^a^ | **Mean difference in trajectory (95% CI) comparing depression at 18 weeks only with the reference trajectory (% or %/yr)** ^b^ | **Mean difference in trajectory (95% CI) comparing depression at 32 weeks only with the reference trajectory (% or %/yr)** ^b^ | **Mean difference in trajectory (95% CI) comparing depression at 18 and 32 weeks with the reference trajectory (% or %/yr)** ^b^ |
| --- | --- | --- | --- | --- | --- |
| **Insulin** |  |  |  |  |  |
| **Unadjusted** |  |  |  |  |  |
| Birth (mu/l or %) | 1.00 (0.90,1.10)  0.04 (0.03,0.06)  0.13 (0.11,0.15)  -0.15 (-0.19,-0.11)  1.75 (1.67,1.84) | 1.08 (0.99,1.18) | -8.47 (-47.50,30.55)  2.05 (-3.91,8.00)  -2.14 (-8.16,3.88)  5.89 (-7.22,19.00)  14.50 (-17.62,46.62) | 6.35 (-28.02,40.73)  -0.56 (-5.32,4.20)  -3.96 (-9.81,1.89)  7.42 (-5.61,20.46)  -1.65 (-26.79,23.49) | 4.26 (-29.26,37.78)  -2.07 (-6.87,2.73)  3.18 (-3.53,9.89)  -7.27 (-19.75,5.21)  -16.92 (-40.13,6.29) |
| Change 0-9yr (mu/l/yr or %/yr) |  | 0.05 (0.04,0.07) |  |  |  |
| Change 9-15yr (mu/l/yr or %/yr)  Change 15-18yr (mu/l/yr or %/yr) |  | 0.14 (0.12,0.16)  -0.13 (-0.17,-0.09) |  |  |  |
| Age 18yr (mu/l or %) |  | 1.98 (1.89,2.06) |  |  |  |
| **Adjusted** |  |  |  |  |  |
| Birth (mu/l or %) | 1.05 (0.77,1.32)  0.02 (-0.02,0.06)  0.18 (0.13,0.24)  -0.16 (-0.27,-0.05)  1.82 (1.59,2.05) | 1.17 (0.89,1.44) | -8.23 (-47.25,30.78)  1.86 (-4.02,7.74)  -1.40 (-7.48,4.68)  5.68 (-7.34,18.69)  17.45 (-15.03,49.92) | 4.98 (-28.32,38.28)  -0.61 (-5.27,4.06)  -3.97 (-9.85,1.91)  9.24 (-4.01,22.49)  1.61 (-24.14,27.37) | 14.00 (-22.85,50.85)  -3.06 (-7.79,1.67)  2.66 (-3.99,9.30)  -5.79 (-18.31,6.73)  -15.65 (-38.77,7.47) |
| Change 0-9yr (mu/l/yr or %/yr) |  | 0.02 (-0.02,0.07) |  |  |  |
| Change 9-15yr (mu/l/yr or %/yr)  Change 15-18yr (mu/l/yr or %/yr) |  | 0.18 (0.13,0.24)  -0.14 (-0.25,-0.03) |  |  |  |
| Age 18yr (mu/l or %) |  | 2.05 (1.82,2.28) |  |  |  |
| **Triglyceride** |  |  |  |  |  |
| **Unadjusted** |  |  |  |  |  |
| Birth (mmol/l or %) | -0.71 (-0.73,-0.69)  0.08 (0.08,0.09)  -0.04 (-0.04,-0.04)  -0.33 (-0.34,-0.31) | -0.70 (-0.72,-0.68) | 1.36 (-4.96,7.68)  0.06 (-0.90,1.03)  -0.46 (-1.33,0.41)  -2.20 (-7.98,3.59) | 0.21 (-5.58,6.00)  -0.09 (-0.95,0.77)  -0.35 (-1.11,0.42)  -3.67 (-8.77,1.43) | -0.49 (-6.46,5.49)  -0.23 (-1.15,0.68)  -0.22 (-1.07,0.63)  -4.44 (-10.11,1.23) |
| Change 0-9yr (mmol/l/yr or %/yr) |  | 0.09 (0.09,0.09) |  |  |  |
| Change 9-18yr (mmol/l/yr or %/yr) |  | -0.04 (-0.05,-0.04) |  |  |  |
| Age 18yr (mmol/l or %) |  | -0.29 (-0.31,-0.27) |  |  |  |
| **Adjusted** |  |  |  |  |  |
| Birth (mmol/l or %) | -0.66 (-0.72,-0.60)  0.08 (0.07,0.09)  -0.04 (-0.05,-0.03)  -0.31 (-0.37,-0.26) | -0.66 (-0.72,-0.60) | -0.67 (-6.76,5.42)  0.24 (-0.72,1.20)  -0.61 (-1.48,0.26)  -3.93 (-9.63,1.77) | 0.90 (-4.82,6.62)  -0.14 (-0.99,0.72)  -0.46 (-1.22,0.31)  -4.38 (-9.45,0.70) | -0.63 (-6.50,5.24)  -0.18 (-1.09,0.73)  -0.41 (-1.26,0.44)  -5.77 (-11.39,-0.14) |
| Change 0-9yr (mmol/l/yr or %/yr) |  | 0.09 (0.08,0.10) |  |  |  |
| Change 9-18yr (mmol/l/yr or %/yr) |  | -0.05 (-0.05,-0.04) |  |  |  |
| Age 18yr (mmol/l or %) |  | -0.28 (-0.33,-0.22) |  |  |  |

CI, confidence interval; mmol/l, millimole per litre; mmol/l/year, millimoles per litre per year; %/yr, percentage per year

^a^ Insulin and triglyceride were transformed using the natural log. All predicted mean values (mmol/l) and rates of change per year (mmol/l/yr) for the reference categories are on the log scale.

^b^ The difference between groups is back transformed from the log scale for ease of interpretation and is interpreted as the percentage difference in the mean level in original units comparing each category with the reference or percentage difference in change in original units per year (%/yr) comparing each category with the reference.

**Supplemental Table S23 Mean trajectories of HDL-C and non-HDL-c estimated from multilevel models, by maternal depression during pregnancy**

|  | **Mean trajectory (95% CI) in males**  **(no depression during pregnancy) (reference)** | **Mean trajectory (95% CI) in females**  **(no depression during pregnancy) (reference)** | **Mean difference in trajectory (95% CI) comparing depression at 18 weeks only with the reference trajectory** | **Mean difference in trajectory (95% CI) comparing depression at 32 weeks only with the reference trajectory** | **Mean difference in trajectory (95% CI) comparing depression at 18 and 32 weeks with the reference trajectory** |
| --- | --- | --- | --- | --- | --- |
| **HDL-c** |  |  |  |  |  |
| **Unadjusted** |  |  |  |  |  |
| Birth (mmol/l) | 0.50 (0.49,0.52)  0.15 (0.15,0.15)  -0.04 (-0.04,-0.03)  1.15 (1.14,1.16) | 0.55 (0.54,0.56) | -0.02 (-0.05,0.02)  0.002 (-0.01,0.01)  0.001 (-0.003,0.01)  0.01 (-0.03,0.05) | -0.04 (-0.07,-0.01)  0.01 (0.001,0.01)  -0.005 (-0.01,-0.001)  -0.03 (-0.07,0.003) | -0.001 (-0.04,0.03)  0.0001 (-0.01,0.01)  -0.001 (-0.01,0.003)  -0.02 (-0.06,0.03) |
| Change 0-7yr (mmol/l/yr) |  | 0.13 (0.13,0.13) |  |  |  |
| Change 7-18yr (mmol/l/yr) |  | -0.01 (-0.01,-0.01) |  |  |  |
| Age 18yr (mmol/l) |  | 1.33 (1.32,1.35) |  |  |  |
| **Adjusted** |  |  |  |  |  |
| Birth (mmol/l) | 0.49 (0.46,0.53)  0.15 (0.14,0.16)  -0.04 (-0.04,-0.03)  1.01 (0.98,1.05)) | 0.54 (0.51,0.58) | -0.01 (-0.04,0.03)  0.001 (-0.01,0.01)  0.002 (-0.002,0.01)  0.03 (-0.02,0.07) | -0.04 (-0.07,-0.01)  0.01 (0.001,0.01)  -0.004 (-0.01,0.0002)  -0.02 (-0.06,0.01) | 0.004 (-0.03,0.04)  -0.001 (-0.01,0.01)  -0.00004 (-0.004,0.004)  -0.001 (-0.04,0.04) |
| Change 0-7yr (mmol/l/yr) |  | 0.13 (0.13,0.14) |  |  |  |
| Change 7-18yr (mmol/l/yr) |  | -0.01 (-0.01,-0.01) |  |  |  |
| Age 18yr (mmol/l) |  | 1.35 (1.31,1.39) |  |  |  |
| **Non-HDL-c** |  |  |  |  |  |
| **Unadjusted** |  |  |  |  |  |
| Birth (mmol/l) | 1.22 (1.20,1.25)  0.19 (0.19,0.19)  -0.07 (-0.08,-0.07)  2.28 (2.24,2.31) | 1.30 (1.27,1.33) | -0.06 (-0.17,0.05)  0.01 (-0.01,0.03)  0.01 (-0.01,0.03)  0.11 (-0.03,0.24) | 0.07 (-0.03,0.17)  -0.01 (-0.03,0.00)  -0.002 (-0.02,0.01)  -0.07 (-0.19,0.06) | -0.04 (-0.15,0.07)  0.005 (-0.01,0.02)  0.005 (-0.01,0.02)  0.05 (-0.09,0.19) |
| Change 0-9yr (mmol/l/yr) |  | 0.20 (0.20,0.21) |  |  |  |
| Change 9-18yr (mmol/l/yr) |  | -0.07 (-0.08,-0.07) |  |  |  |
| Age 18yr (mmol/l) |  | 2.48 (2.45,2.51) |  |  |  |
| **Adjusted** |  |  |  |  |  |
| Birth (mmol/l) | 1.20 (1.12,1.28)  0.19 (0.18,0.20)  -0.07 (-0.08,-0.06)  2.28 (2.19,2.36) | 1.27 (1.19,1.35) | -0.08 (-0.19,0.04)  0.01 (-0.01,0.03)  0.01 (-0.01,0.02)  0.09 (-0.05,0.22) | 0.07 (-0.03,0.18)  -0.01 (-0.03,0.00)  -0.003 (-0.02,0.01)  -0.08 (-0.20,0.04) | -0.03 (-0.15,0.08)  0.004 (-0.01,0.02)  0.004 (-0.01,0.02)  0.04 (-0.10,0.18) |
| Change 0-9yr (mmol/l/yr) |  | 0.21 (0.20,0.22) |  |  |  |
| Change 9-18yr (mmol/l/yr) |  | -0.07 (-0.08,-0.06) |  |  |  |
| Age 18yr (mmol/l) |  | 2.48 (2.39,2.57) |  |  |  |

CI, confidence interval; mmol/l, millimole per litre; mmol/l, millimole per litre per year.

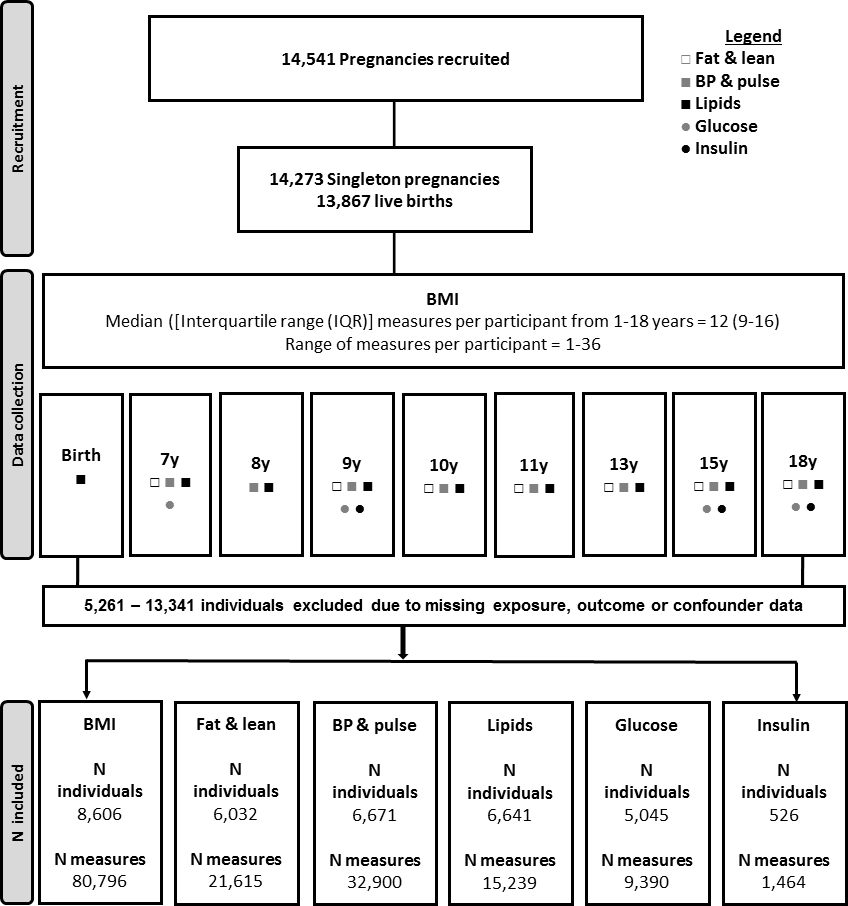

**Supplemental Figure S1 Flow diagram of study design**

**Legend:** BP, blood pressure; BMI, body mass index; y, years of age

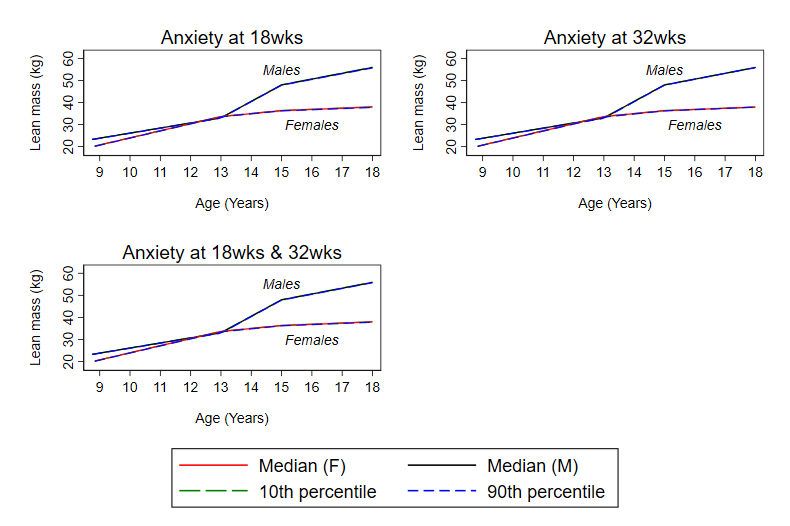

**Supplemental Figure S2 Mean predicted confounder adjusted trajectories of lean mass (9 to 18 years), by continuous maternal anxiety levels during pregnancy**

**Legend:** Trajectories are adjusted for maternal age at birth of offspring, parental household socioeconomic position, parity, maternal pre-pregnancy BMI and maternal smoking during pregnancy. Median, 10^th^ and 90^th^ percentile = 1, 4 and 9 at 18 weeks. Median, 10^th^ and 90^th^ percentile = 1, 4, 9 at 32 weeks. Mean of the 18 and 32 week measures used for analyses of anxiety at 18 weeks and 32 weeks; median, 10^th^ and 90^th^ percentile = 1, 4, 9 in this analysis.

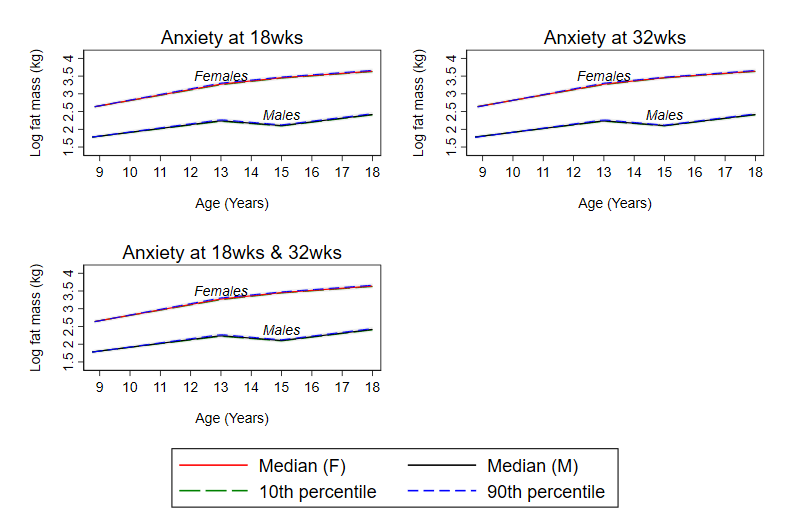

**Supplemental Figure S3 Mean predicted confounder adjusted trajectories of log fat mass (9 to 18 years), by continuous maternal anxiety levels during pregnancy**

**Legend:** Trajectories are adjusted for maternal age at birth of offspring, parental household socioeconomic position, parity, maternal pre-pregnancy BMI and maternal smoking during pregnancy. Median, 10^th^ and 90^th^ percentile = 1, 4 and 9 at 18 weeks. Median, 10^th^ and 90^th^ percentile = 1, 4, 10 at 32 weeks. Mean of the 18 and 32 week measures used for analyses of anxiety at 18 weeks and 32 weeks; median, 10^th^ and 90^th^ percentile = 1, 4, 9 in this analysis.

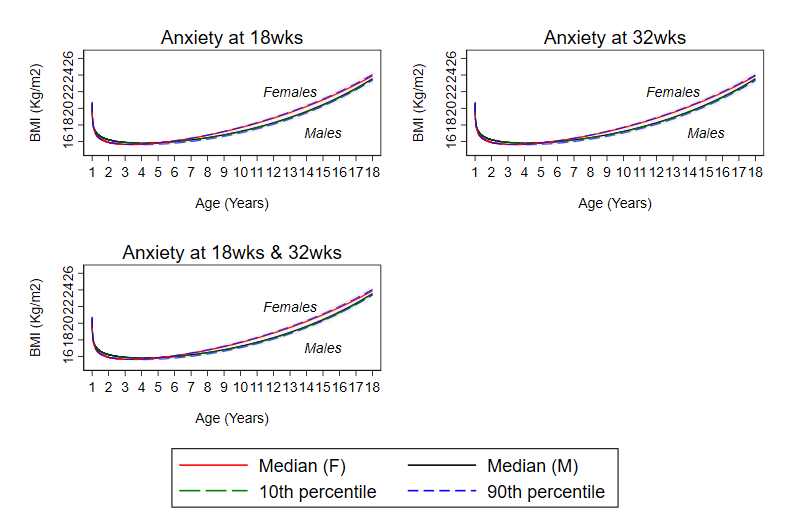

**Supplemental Figure S4 Mean predicted confounder adjusted trajectories of log BMI (1 to 18 years), by continuous maternal anxiety levels during pregnancy**

**Legend:** Trajectories are adjusted for maternal age at birth of offspring, parental household socioeconomic position, parity, maternal pre-pregnancy BMI and maternal smoking during pregnancy. Median, 10^th^ and 90^th^ percentile = 1, 4 and 9 at 18 weeks. Median, 10^th^ and 90^th^ percentile = 1, 4, 9 at 32 weeks. Mean of the 18 and 32 week measures used for analyses of anxiety at 18 weeks and 32 weeks; median, 10^th^ and 90^th^ percentile = 1, 4, 9 in this analysis.

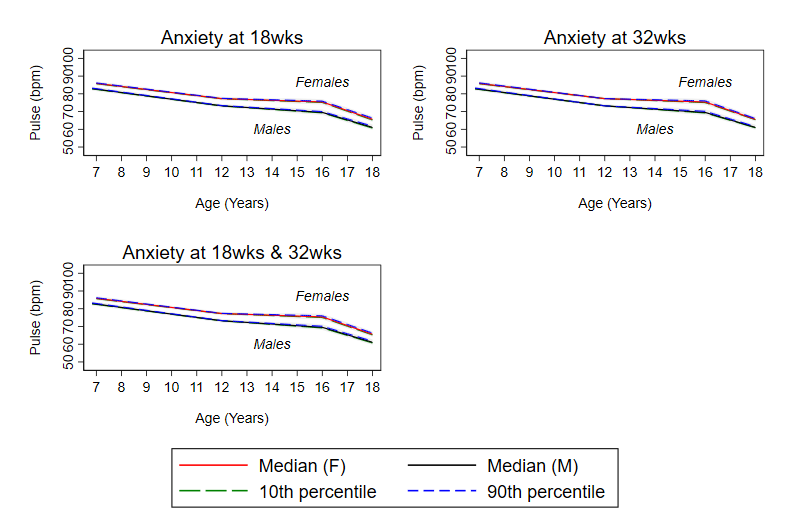

**Supplemental Figure S5 Mean predicted confounder adjusted trajectories of pulse rate from 7 to 18 years by continuous maternal anxiety levels during pregnancy**

**Legend:** Trajectories are adjusted for maternal age at birth of offspring, parental household socioeconomic position, parity, maternal pre-pregnancy BMI and maternal smoking during pregnancy. Median, 10^th^ and 90^th^ percentile = 1, 4 and 9 at 18 weeks. Median, 10^th^ and 90^th^ percentile = 1, 4, 10 at 32 weeks. Mean of the 18 and 32 week measures used for analyses of anxiety at 18 weeks and 32 weeks; median, 10^th^ and 90^th^ percentile = 1, 4, 9 in this analysis.

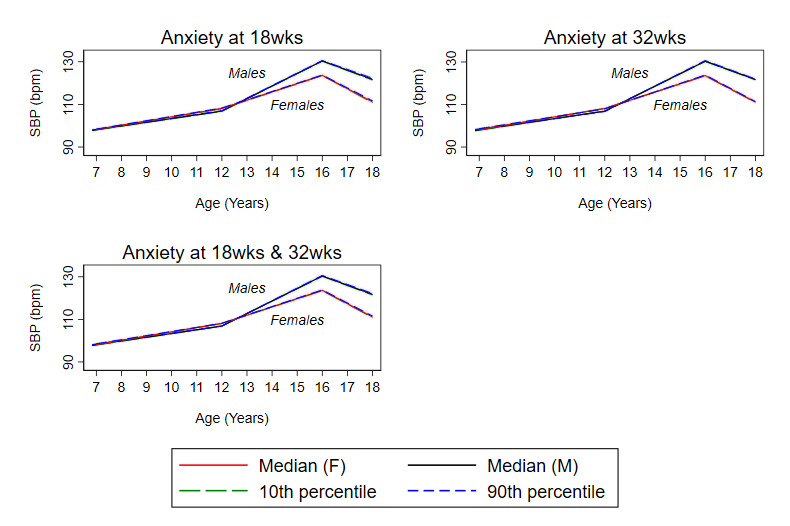

**Supplemental Figure S6 Mean predicted confounder adjusted trajectories of SBP from 7 to 18 years by continuous maternal anxiety levels during pregnancy**

**Legend:** Trajectories are adjusted for maternal age at birth of offspring, parental household socioeconomic position, parity, maternal pre-pregnancy BMI and maternal smoking during pregnancy. Median, 10^th^ and 90^th^ percentile = 1, 4 and 9 at 18 weeks. Median, 10^th^ and 90^th^ percentile = 1, 4, 10 at 32 weeks. Mean of the 18 and 32 week measures used for analyses of anxiety at 18 weeks and 32 weeks; median, 10^th^ and 90^th^ percentile = 1, 4, 9 in this analysis.

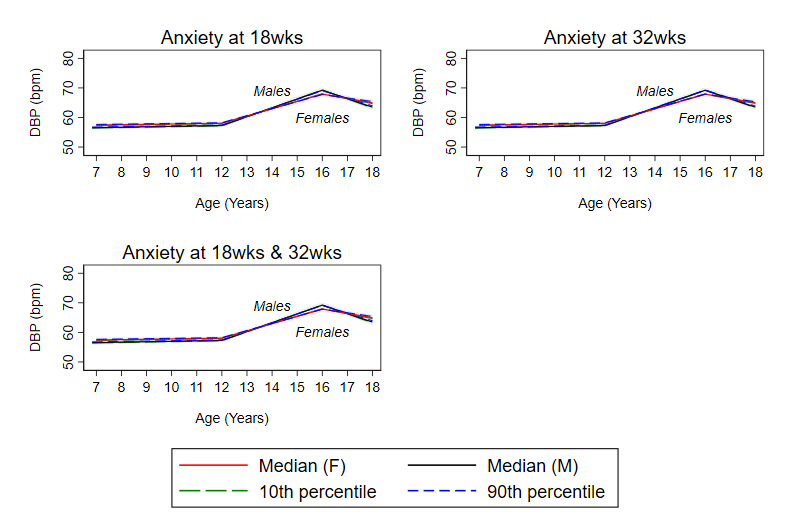

**Supplemental Figure S7 Mean predicted confounder adjusted trajectories of DBP from 7 to 18 years by continuous maternal anxiety levels during pregnancy**

**Legend:** Trajectories are adjusted for maternal age at birth of offspring, parental household socioeconomic position, parity, maternal pre-pregnancy BMI and maternal smoking during pregnancy. Median, 10^th^ and 90^th^ percentile = 1, 4 and 9 at 18 weeks. Median, 10^th^ and 90^th^ percentile = 1, 4, 10 at 32 weeks. Mean of the 18 and 32 week measures used for analyses of anxiety at 18 weeks and 32 weeks; median, 10^th^ and 90^th^ percentile = 1, 4, 9 in this analysis.

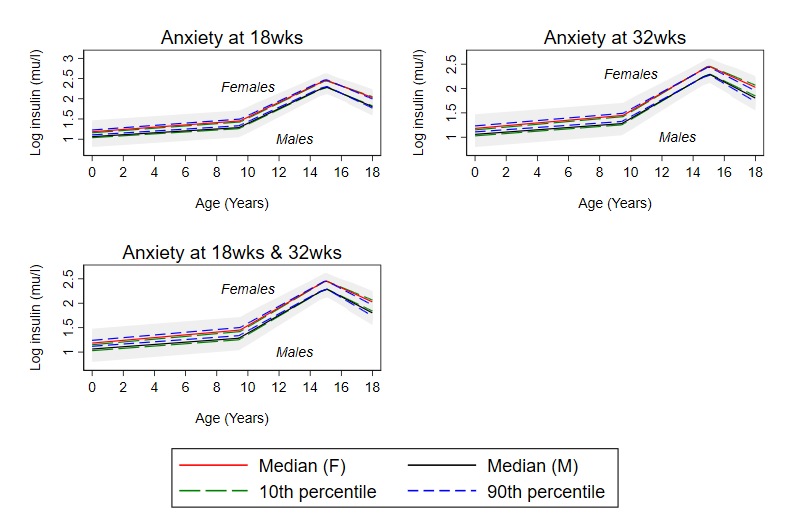

**Supplemental Figure S8 Mean predicted confounder adjusted trajectories of log insulin from (birth to 18 years) by continuous maternal anxiety levels during pregnancy**

**Legend:** Trajectories are adjusted for maternal age at birth of offspring, parental household socioeconomic position, parity, maternal pre-pregnancy BMI and maternal smoking during pregnancy. Median, 10^th^ and 90^th^ percentile = 1, 4 and 9 at 18 weeks. Median, 10^th^ and 90^th^ percentile = 1, 4, 9 at 32 weeks. Mean of the 18 and 32 week measures used for analyses of anxiety at 18 weeks and 32 weeks; median, 10^th^ and 90^th^ percentile = 1, 4, 9 in this analysis.

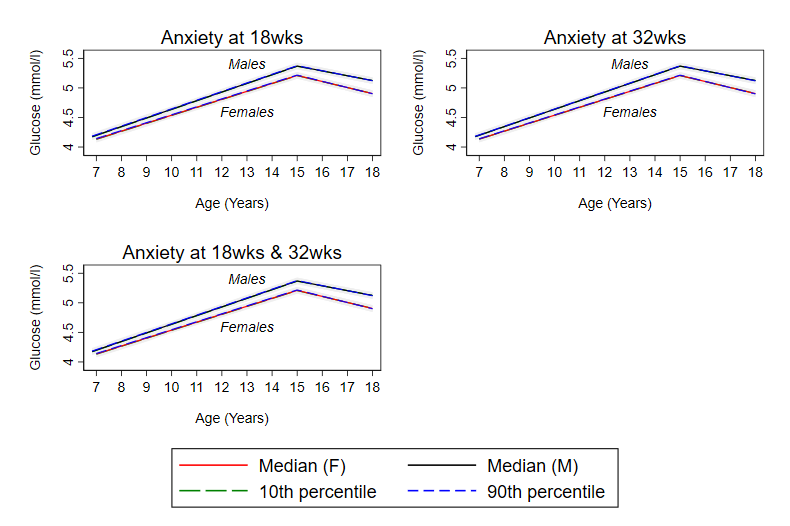

**Supplemental Figure S9 Mean predicted confounder adjusted trajectories of glucose (7 to18 years), by continuous maternal anxiety levels during pregnancy**

**Legend:** Trajectories are adjusted for maternal age at birth of offspring, parental household socioeconomic position, parity, maternal pre-pregnancy BMI and maternal smoking during pregnancy. Median, 10^th^ and 90^th^ percentile = 1, 4 and 9 at 18 weeks. Median, 10^th^ and 90^th^ percentile = 1, 4, 10 at 32 weeks. Mean of the 18 and 32 week measures used for analyses of anxiety at 18 weeks and 32 weeks; median, 10^th^ and 90^th^ percentile = 1, 4, 9 in this analysis.

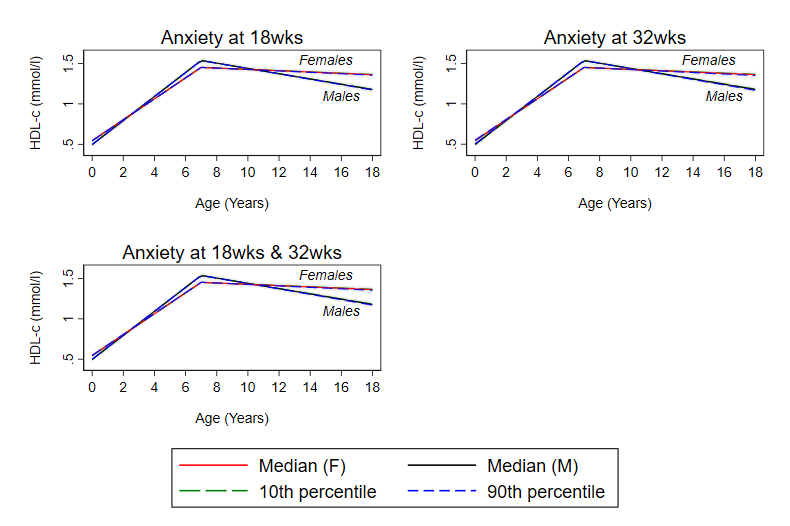

**Supplemental Figure S10 Mean predicted confounder adjusted trajectories of HDL-c (birth to18 years), by continuous maternal anxiety levels during pregnancy**

**Legend:** Trajectories are adjusted for maternal age at birth of offspring, parental household socioeconomic position, parity, maternal pre-pregnancy BMI and maternal smoking during pregnancy. Median, 10^th^ and 90^th^ percentile = 1, 4 and 9 at 18 weeks. Median, 10^th^ and 90^th^ percentile = 1, 4, 10 at 32 weeks. Mean of the 18 and 32 week measures used for analyses of anxiety at 18 weeks and 32 weeks; median, 10^th^ and 90^th^ percentile = 1, 4, 9 in this analysis.

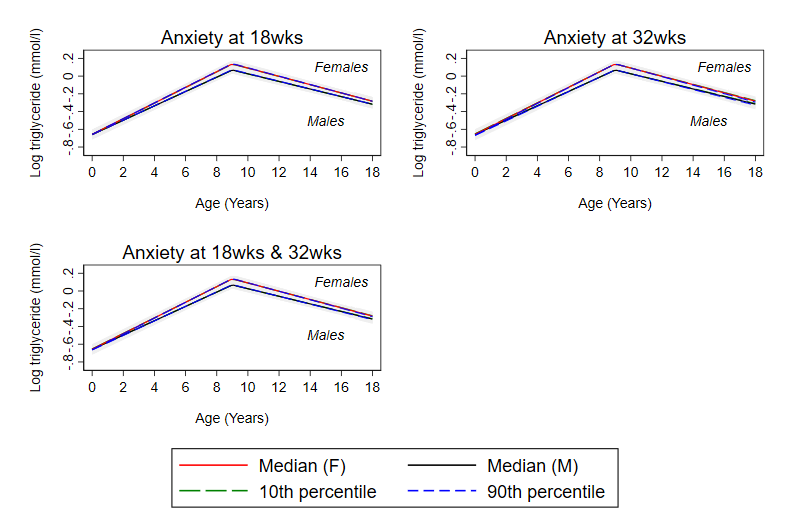

**Supplemental Figure S11 Mean predicted confounder adjusted trajectories of triglyceride (birth to18 years), by continuous maternal anxiety levels during pregnancy**

**Legend:** Trajectories are adjusted for maternal age at birth of offspring, parental household socioeconomic position, parity, maternal pre-pregnancy BMI and maternal smoking during pregnancy. Median, 10^th^ and 90^th^ percentile = 1, 4 and 9 at 18 weeks. Median, 10^th^ and 90^th^ percentile = 1, 4, 10 at 32 weeks. Mean of the 18 and 32 week measures used for analyses of anxiety at 18 weeks and 32 weeks; median, 10^th^ and 90^th^ percentile = 1, 4, 9 in this analysis.

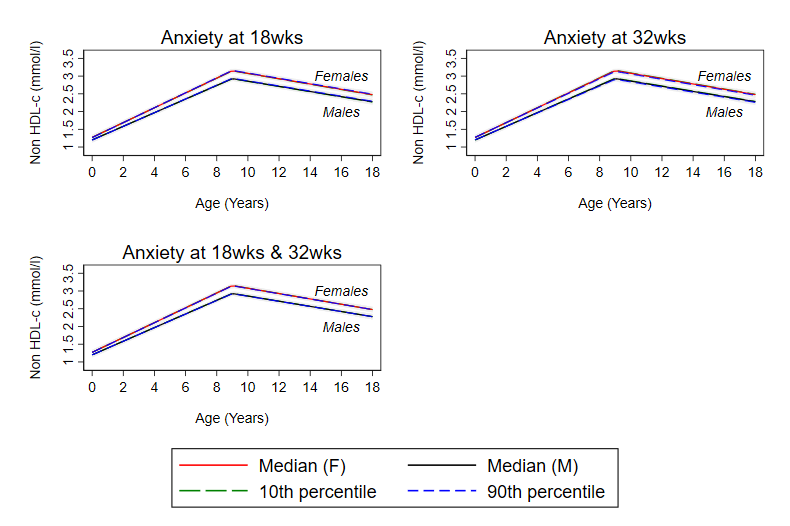

**Supplemental Figure S12 Mean predicted confounder adjusted trajectories of non-HDL-c (birth to18 years), by continuous maternal anxiety levels during pregnancy**

**Legend:** Trajectories are adjusted for maternal age at birth of offspring, parental household socioeconomic position, parity, maternal pre-pregnancy BMI and maternal smoking during pregnancy. Median, 10^th^ and 90^th^ percentile = 1, 4 and 9 at 18 weeks. Median, 10^th^ and 90^th^ percentile = 1, 4, 10 at 32 weeks. Mean of the 18 and 32 week measures used for analyses of anxiety at 18 weeks and 32 weeks; median, 10^th^ and 90^th^ percentile = 1, 4, 9 in this analysis.

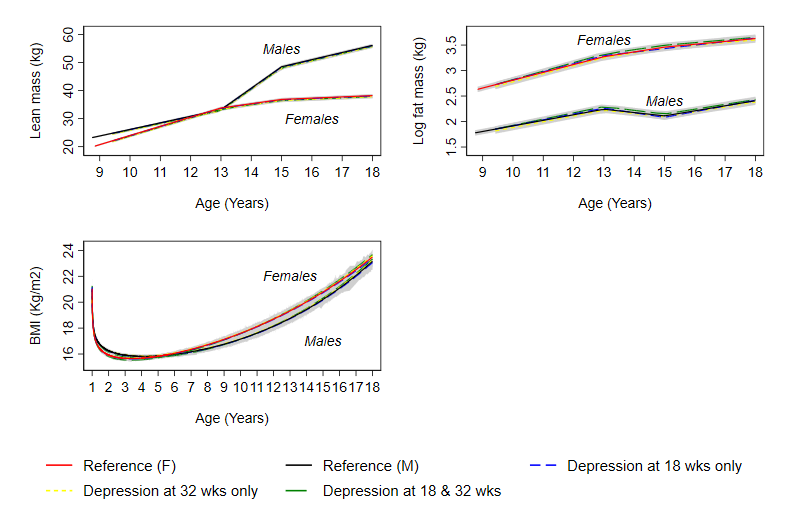

**Supplemental Figure S13 Mean predicted confounder adjusted trajectories of lean mass (9 to 18 years), log fat mass (9 to 18 years) and log BMI (1 to 18 years), by maternal depression levels during pregnancy**

**Legend:** Trajectories are adjusted for maternal age at birth of offspring, parental household socioeconomic position, parity, maternal pre-pregnancy BMI and maternal smoking during pregnancy.

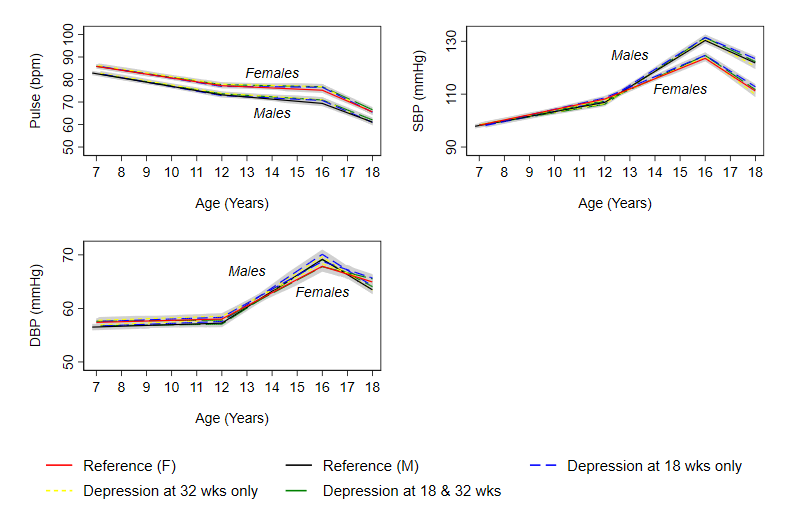
 **Supplemental Figure S14 Mean predicted confounder adjusted trajectories of pulse rate, SBP and DBP from 7 to 18 years, by maternal depression levels during pregnancy**

**Legend:** DBP, diastolic blood pressure; SBP, systolic blood pressure.

Trajectories are adjusted for maternal age at birth of offspring, parental household socioeconomic position, parity, maternal pre-pregnancy BMI and maternal smoking during pregnancy. Depression at 18 and 32 weeks gestation defined as ≥85^th^ percentile of depression score based on Edinburgh Postnatal Depression Scale for the whole cohort at each time point separately ((≥85^th^ percentile = 13 at both 18 and 32 weeks gestation).

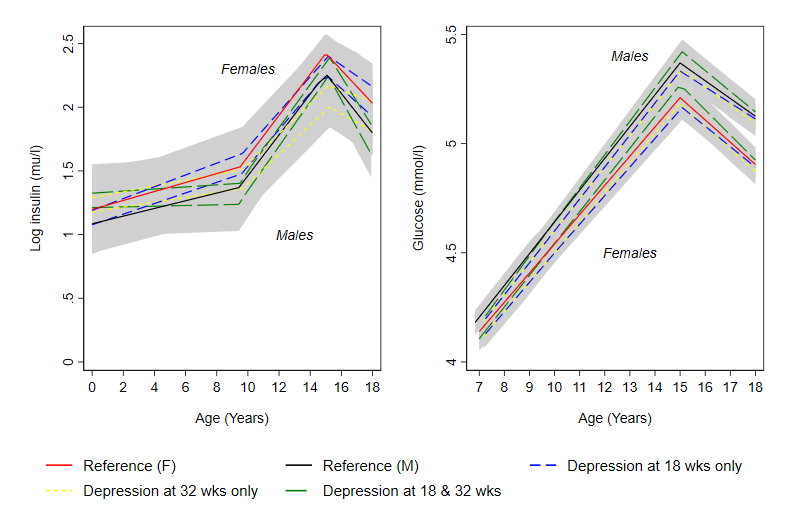
 **Supplemental Figure S15 Mean predicted confounder adjusted trajectories of log insulin (birth to 18 years) and glucose (7 to 18 years), by maternal depression levels during pregnancy**

**Legend:** Trajectories are adjusted for maternal age at birth of offspring, parental household socioeconomic position, parity, maternal pre-pregnancy BMI and maternal smoking during pregnancy. Depression at 18 and 32 weeks gestation defined as ≥85^th^ percentile of depression score based on Edinburgh Postnatal Depression Scale for the whole cohort at each time point separately ((≥85^th^ percentile = 13 at both 18 and 32 weeks gestation).

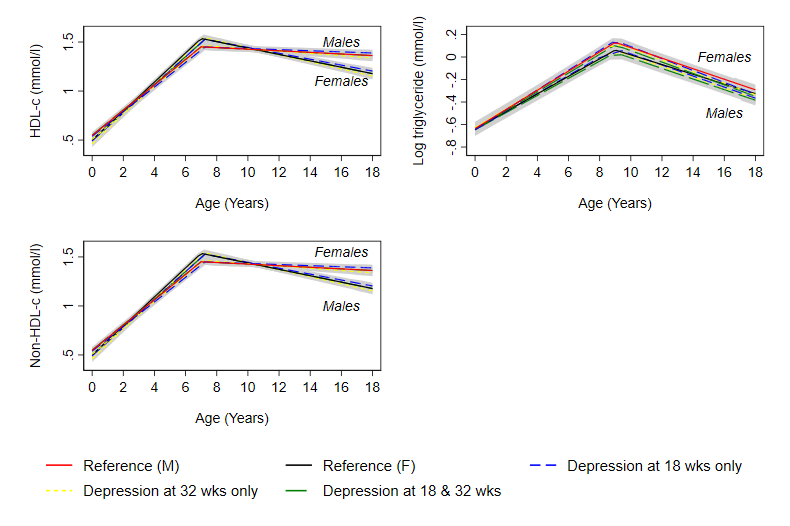
 **Supplemental Figure S16 Mean predicted confounder adjusted trajectories of HDL-c, log triglyceride and non-HDL-c from birth to 18 years, by maternal depression levels during pregnancy**

**Legend:** Trajectories are adjusted for maternal age at birth of offspring, parental household socioeconomic position, parity, maternal pre-pregnancy BMI and maternal smoking during pregnancy. Depression at 18 and 32 weeks gestation defined as ≥85^th^ percentile of depression score based on Edinburgh Postnatal Depression Scale for the whole cohort at each time point separately ((≥85^th^ percentile = 13 at both 18 and 32 weeks gestation).

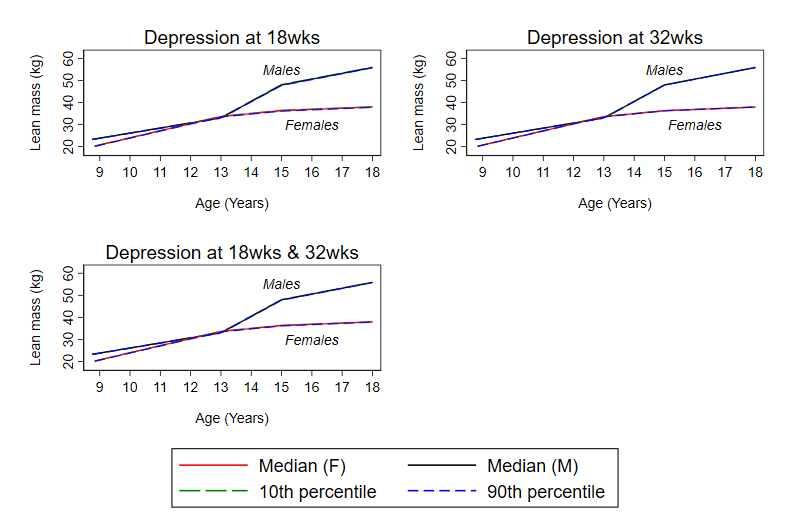

**Supplemental Figure S17 Mean predicted confounder adjusted trajectories of lean mass (9 to 18 years), by continuous maternal depression levels during pregnancy**

**Legend:** Trajectories are adjusted for maternal age at birth of offspring, parental household socioeconomic position, parity, maternal pre-pregnancy BMI and maternal smoking during pregnancy. Median, 10^th^ and 90^th^ percentile = 1, 6 and 12 at 18 weeks. Median, 10^th^ and 90^th^ percentile = 1, 6, 13 at 32 weeks. Mean of the 18 and 32 week measures used for analyses of anxiety at 18 weeks and 32 weeks; median, 10^th^ and 90^th^ percentile = 1.5, 5.5, 12 in this analysis.

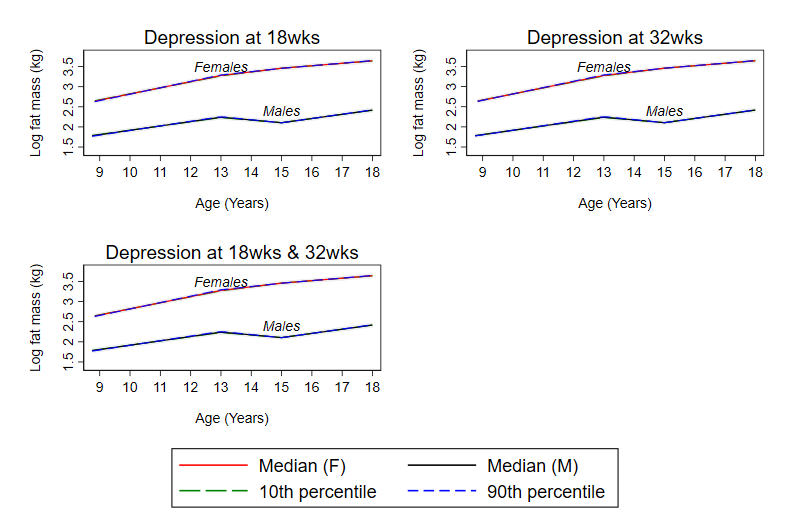

**Supplemental Figure S18 Mean predicted confounder adjusted trajectories of log fat mass (9 to 18 years), by continuous maternal depression levels during pregnancy**

**Legend:** Trajectories are adjusted for maternal age at birth of offspring, parental household socioeconomic position, parity, maternal pre-pregnancy BMI and maternal smoking during pregnancy. Median, 10^th^ and 90^th^ percentile = 1, 6 and 12 at 18 weeks. Median, 10^th^ and 90^th^ percentile = 1, 6, 13 at 32 weeks. Mean of the 18 and 32 week measures used for analyses of anxiety at 18 weeks and 32 weeks; median, 10^th^ and 90^th^ percentile = 1.5, 5.5, 12 in this analysis.

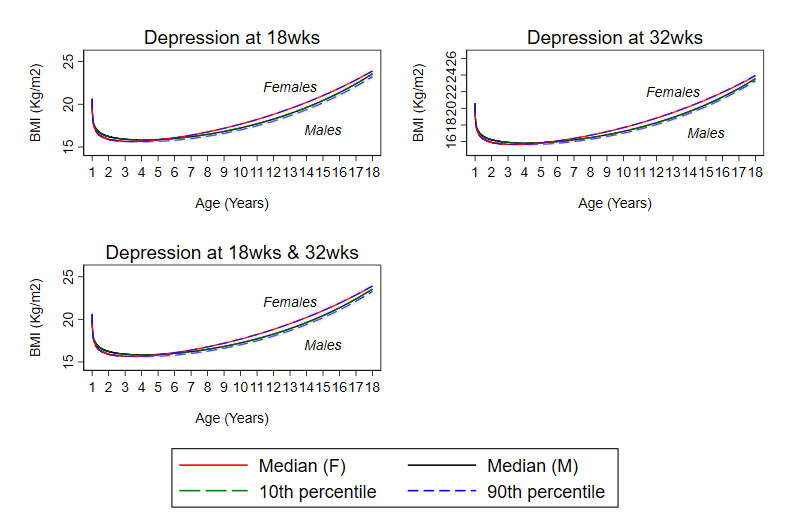

**Supplemental Figure S19 Mean predicted confounder adjusted trajectories of log BMI (1 to 18 years), by continuous maternal depression levels during pregnancy**

**Legend:** Trajectories are adjusted for maternal age at birth of offspring, parental household socioeconomic position, parity, maternal pre-pregnancy BMI and maternal smoking during pregnancy. Median, 10^th^ and 90^th^ percentile = 1, 6 and 13 at 18 weeks. Median, 10^th^ and 90^th^ percentile = 1, 6, 13 at 32 weeks. Mean of the 18 and 32 week measures used for analyses of anxiety at 18 weeks and 32 weeks; median, 10^th^ and 90^th^ percentile = 1.5, 5.5, 12 in this analysis.

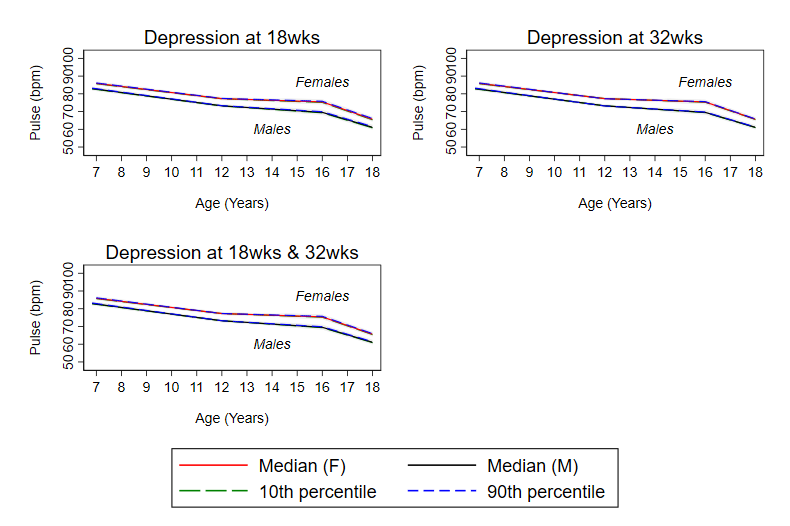

**Supplemental Figure S20 Mean predicted confounder adjusted trajectories of pulse rate from 7 to 18 years by continuous maternal depression levels during pregnancy**

**Legend:** Trajectories are adjusted for maternal age at birth of offspring, parental household socioeconomic position, parity, maternal pre-pregnancy BMI and maternal smoking during pregnancy. Median, 10^th^ and 90^th^ percentile = 1, 6 and 13 at 18 weeks. Median, 10^th^ and 90^th^ percentile = 1, 6, 13 at 32 weeks. Mean of the 18 and 32 week measures used for analyses of anxiety at 18 weeks and 32 weeks; median, 10^th^ and 90^th^ percentile = 1.5, 5.5, 12 in this analysis.

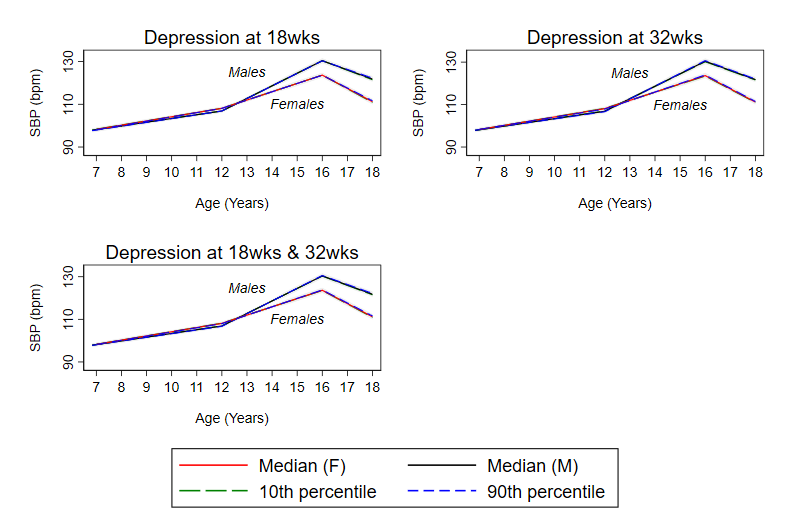

**Supplemental Figure S21 Mean predicted confounder adjusted trajectories of SBP from 7 to 18 years by continuous maternal depression levels during pregnancy**

**Legend:** Trajectories are adjusted for maternal age at birth of offspring, parental household socioeconomic position, parity, maternal pre-pregnancy BMI and maternal smoking during pregnancy. Median, 10^th^ and 90^th^ percentile = 1, 6 and 13 at 18 weeks. Median, 10^th^ and 90^th^ percentile = 1, 6, 13 at 32 weeks. Mean of the 18 and 32 week measures used for analyses of anxiety at 18 weeks and 32 weeks; median, 10^th^ and 90^th^ percentile = 1.5, 5.5, 12 in this analysis.

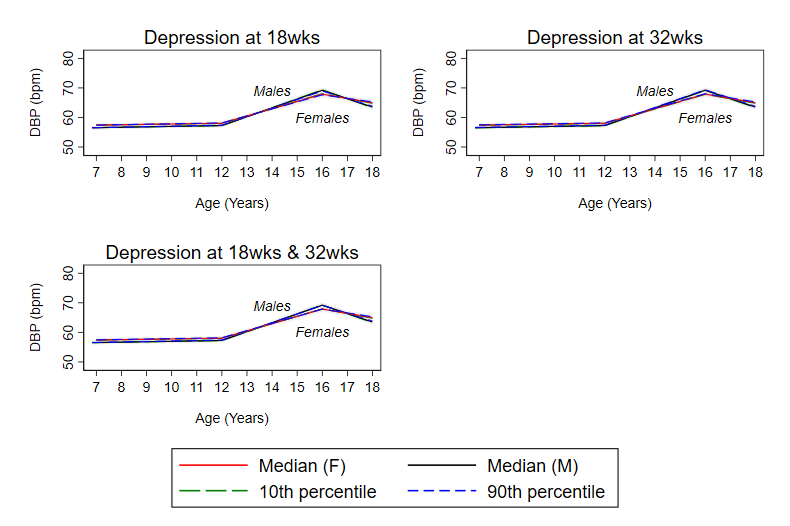

**Supplemental Figure S22 Mean predicted confounder adjusted trajectories of DBP from 7 to 18 years by continuous maternal depression levels during pregnancy**

**Legend:** Trajectories are adjusted for maternal age at birth of offspring, parental household socioeconomic position, parity, maternal pre-pregnancy BMI and maternal smoking during pregnancy. Median, 10^th^ and 90^th^ percentile = 1, 6 and 13 at 18 weeks. Median, 10^th^ and 90^th^ percentile = 1, 6, 13 at 32 weeks. Mean of the 18 and 32 week measures used for analyses of anxiety at 18 weeks and 32 weeks; median, 10^th^ and 90^th^ percentile = 1.5, 5.5, 12 in this analysis.

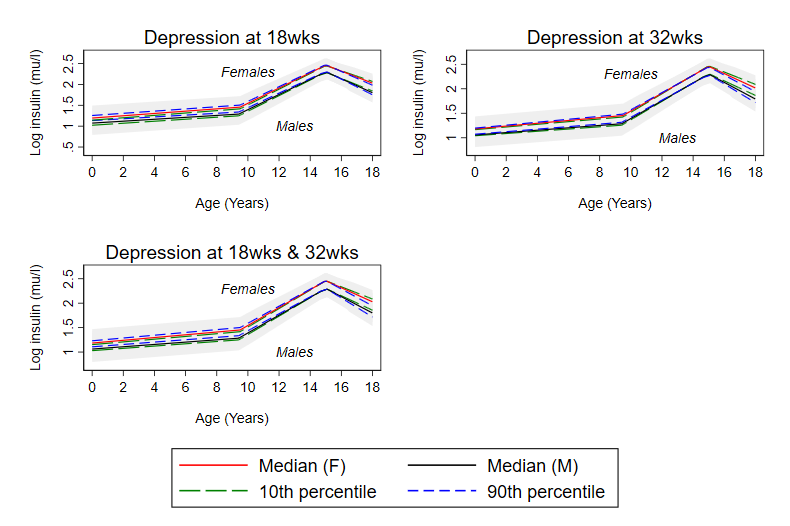

**Supplemental Figure S23 Mean predicted confounder adjusted trajectories of log insulin from (birth to 18 years) by continuous maternal depression levels during pregnancy**

**Legend:** Trajectories are adjusted for maternal age at birth of offspring, parental household socioeconomic position, parity, maternal pre-pregnancy BMI and maternal smoking during pregnancy. Median, 10^th^ and 90^th^ percentile = 1, 5 and 13 at 18 weeks. Median, 10^th^ and 90^th^ percentile = 0, 6, 13 at 32 weeks. Mean of the 18 and 32 week measures used for analyses of anxiety at 18 weeks and 32 weeks; median, 10^th^ and 90^th^ percentile = 1, 5.5, 12 in this analysis.

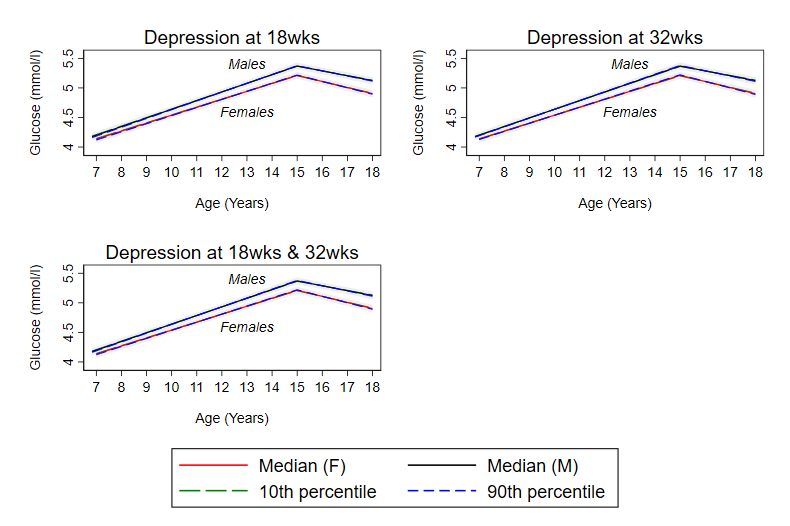

**Supplemental Figure S24 Mean predicted confounder adjusted trajectories of glucose (7 to18 years), by continuous maternal depression levels during pregnancy**

**Legend:** Trajectories are adjusted for maternal age at birth of offspring, parental household socioeconomic position, parity, maternal pre-pregnancy BMI and maternal smoking during pregnancy. Median, 10^th^ and 90^th^ percentile = 1, 6 and 13 at 18 weeks. Median, 10^th^ and 90^th^ percentile = 1, 6, 13 at 32 weeks. Mean of the 18 and 32 week measures used for analyses of anxiety at 18 weeks and 32 weeks; median, 10^th^ and 90^th^ percentile = 1.5, 5.5, 12 in this analysis.

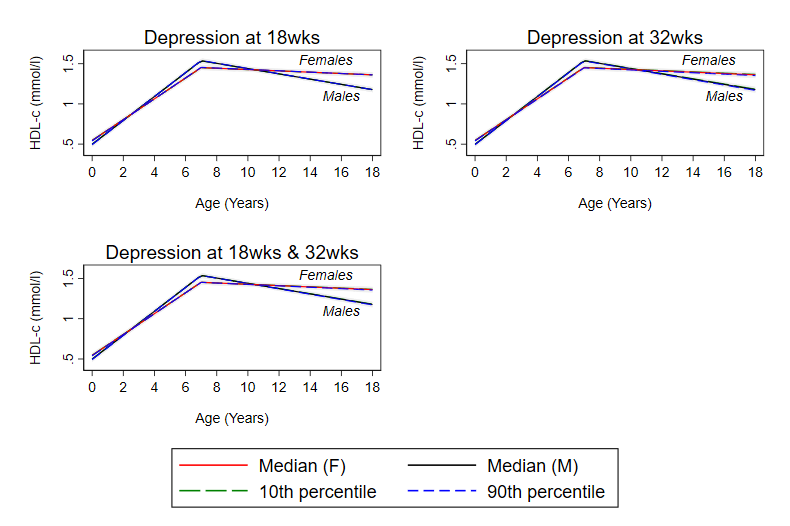

**Supplemental Figure S25 Mean predicted confounder adjusted trajectories of HDL-c (birth to18 years), by continuous maternal depression levels during pregnancy**

**Legend:** Trajectories are adjusted for maternal age at birth of offspring, parental household socioeconomic position, parity, maternal pre-pregnancy BMI and maternal smoking during pregnancy. Median, 10^th^ and 90^th^ percentile = 1, 6 and 13 at 18 weeks. Median, 10^th^ and 90^th^ percentile = 1, 6, 13 at 32 weeks. Mean of the 18 and 32 week measures used for analyses of anxiety at 18 weeks and 32 weeks; median, 10^th^ and 90^th^ percentile = 1.5, 5.5, 12.5 in this analysis.

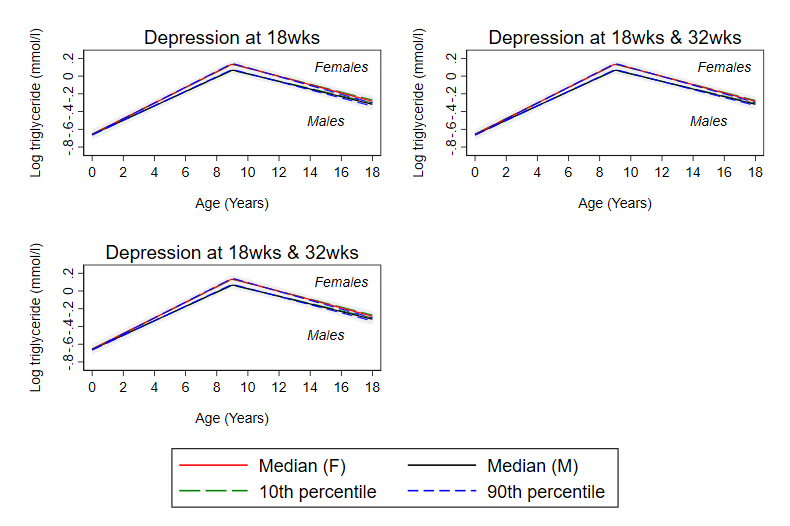

**Supplemental Figure S26 Mean predicted confounder adjusted trajectories of triglyceride (birth to18 years), by continuous maternal depression levels during pregnancy**

**Legend:** Trajectories are adjusted for maternal age at birth of offspring, parental household socioeconomic position, parity, maternal pre-pregnancy BMI and maternal smoking during pregnancy. Median, 10^th^ and 90^th^ percentile = 1, 6 and 13 at 18 weeks. Median, 10^th^ and 90^th^ percentile = 1, 6, 13 at 32 weeks. Mean of the 18 and 32 week measures used for analyses of anxiety at 18 weeks and 32 weeks; median, 10^th^ and 90^th^ percentile = 1.5, 6, 12.5 in this analysis.

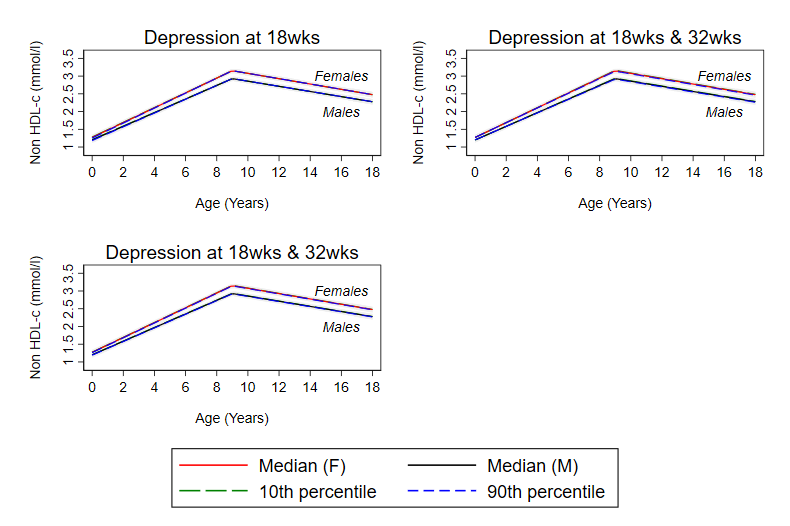

**Supplemental Figure S27 Mean predicted confounder adjusted trajectories of non-HDL-c (birth to18 years), by continuous maternal depression levels during pregnancy**

**Legend:** Trajectories are adjusted for maternal age at birth of offspring, parental household socioeconomic position, parity, maternal pre-pregnancy BMI and maternal smoking during pregnancy. Median, 10^th^ and 90^th^ percentile = 1, 6 and 13 at 18 weeks. Median, 10^th^ and 90^th^ percentile = 1, 6, 13 at 32 weeks. Mean of the 18 and 32 week measures used for analyses of anxiety at 18 weeks and 32 weeks; median, 10^th^ and 90^th^ percentile = 1.5, 6, 12.5 in this analysis.
